# Outbreak risks, cases, and costs of vaccination strategies against wild poliomyelitis in polio-free settings: a modelling study

**DOI:** 10.1101/2023.07.05.23292288

**Authors:** Megan Auzenbergs, Kaja Abbas, Arie Voorman, Corey Peak, Mark Jit, Kathleen M O’Reilly

## Abstract

**Background:** Polio eradication was threatened in 2022 by importation of wild poliovirus serotype 1 into Malawi with subsequent international spread, representing the first wild polio cases in Africa since 2014. Preventing importations and spread of wild poliovirus is critical, and dependent on population immunity provided through routine immunisation and supplementary immunisation activities (SIAs). However, the scale of preventative SIAs has reduced in recent years due to financial constraints.

**Methods:** We developed a mathematical model of polio transmission dynamics to evaluate the probability of an outbreak, expected number of poliomyelitis cases, and the costs associated with vaccination delivery through routine immunisation (RI), outbreak response SIAs (oSIAs) and preventative SIAs (pSIAs). Across varying levels of routine immunisation coverage, we explore three key strategies: RI+oSIAs, RI+oSIAs+annual pSIAs, and RI+oSIAs+biannual pSIAs.

**Results:** The annual pSIA strategy (RI+oSIAs+annual pSIAs) had higher costs but greater probability of no outbreaks: under our model assumptions, annual pSIAs result in 80% probability of no outbreaks when routine immunisation coverage ≥50%. The biannual pSIA strategy (RI+oSIAs+biannual pSIAs) costs less and averts more outbreaks than RI+oSIAs, but RI coverage ≥65% was required to achieve equivalent risk of no outbreaks. The strategy with no pSIAs (RI+oSIAs) had the lowest costs but required ≥75% RI coverage to achieve equivalent risk of no outbreaks.

**Conclusion:** Prioritisation of pSIAs must balance outbreak risk against implementation costs, ideally favouring the smallest manageable outbreak risk compatible with elimination. We infer that there are few short-term risks due to population immunity from RI, but without pSIAs, long-term risks accumulate and can result in outbreaks with potential for international spread. We do not consider the costs of further delaying the eradication timeline or societal implications of outbreaks, both of which emphasise the need for pSIAs.

## Introduction

In 2019, the African Region was certified free from endogenous transmission of wild poliovirus (WPV), with the last case reported in Nigeria in July 2014 (1). However, in early 2022, Malawi and Mozambique reported WPV serotype 1 (WPV1) cases, linked to ongoing circulation in Pakistan (2, 3). The geographic distribution and genetic linkage of these WPV1 cases suggest missed transmission of an unknown geographic extent (2). These WPV1 cases highlight the importance of ensuring high and homogeneous levels of population immunity despite decreasing global incidence and elimination within the African continent (4). Historically, WPV1 outbreaks in Africa had a varied epidemiology— ranging in size from a few cases to explosive outbreaks that resulted in international spread and wide-scale outbreak responses. This variation can be attributed to population exposure to circulating virus and serotype-specific population immunity against poliovirus, among other factors (5). Since 2016, the recommended routine immunisation schedule is with bivalent oral polio vaccine (bOPV) and at least 1 dose of the inactivated polio vaccine (IPV) to provide population immunity. However, variable routine immunisation coverage leads to differential population immunity in different countries. The joint assessment of immunization coverage by UNICEF and WHO (WUENIC) estimated in 2021 routine vaccine coverage in Malawi and Mozambique was 93% and 61%, respectively. Also, low levels of population immunity persist in other areas, such as Liberia, where a recent study demonstrated poliovirus antibody seroprevalence was only 60% against serotype 1 (6).

Further, the number of circulating vaccine derived poliovirus serotype 1 (cVDPV1) outbreaks has increased since 2022, suggesting that population immunity against this serotype has declined in some areas despite a rise in IPV coverage (which is expected to reduce the incidence of paralysis but has limited impact on transmission) (7).

The Global Polio Eradication Initiative (GPEI) is responsible for the coordination of activities to support polio eradication. The activities include surveillance for acute flaccid paralysis (AFP) caused by polio infection, environmental surveillance (ES) for poliovirus, and providing polio vaccinations through both supplementary immunisation activities (SIAs) and routine immunisation (through the Expanded Programme for Immunisation (EPI)). The SIAs aim to vaccinate hard-to-reach children otherwise missed by RI (8). Despite an annual expenditure of hundreds of millions of USD, decision makers within polio eradication often must make difficult decisions in resource allocation. The global budget has remained relatively stable over the last decade with noted decreases in the last three years. Alongside, the frequency of preventative supplementary immunisation activities (pSIAs), which aim to strengthen population immunity and prevent the frequency and impact of outbreaks has decreased in almost all countries in Africa since 2017 (Figure 1) (9). However, the annual costs for vaccination and programmatic activities such as antiviral development and vaccine stockpiling carried out by GPEI have increased considerably (10, 11).

**Figure 1.**
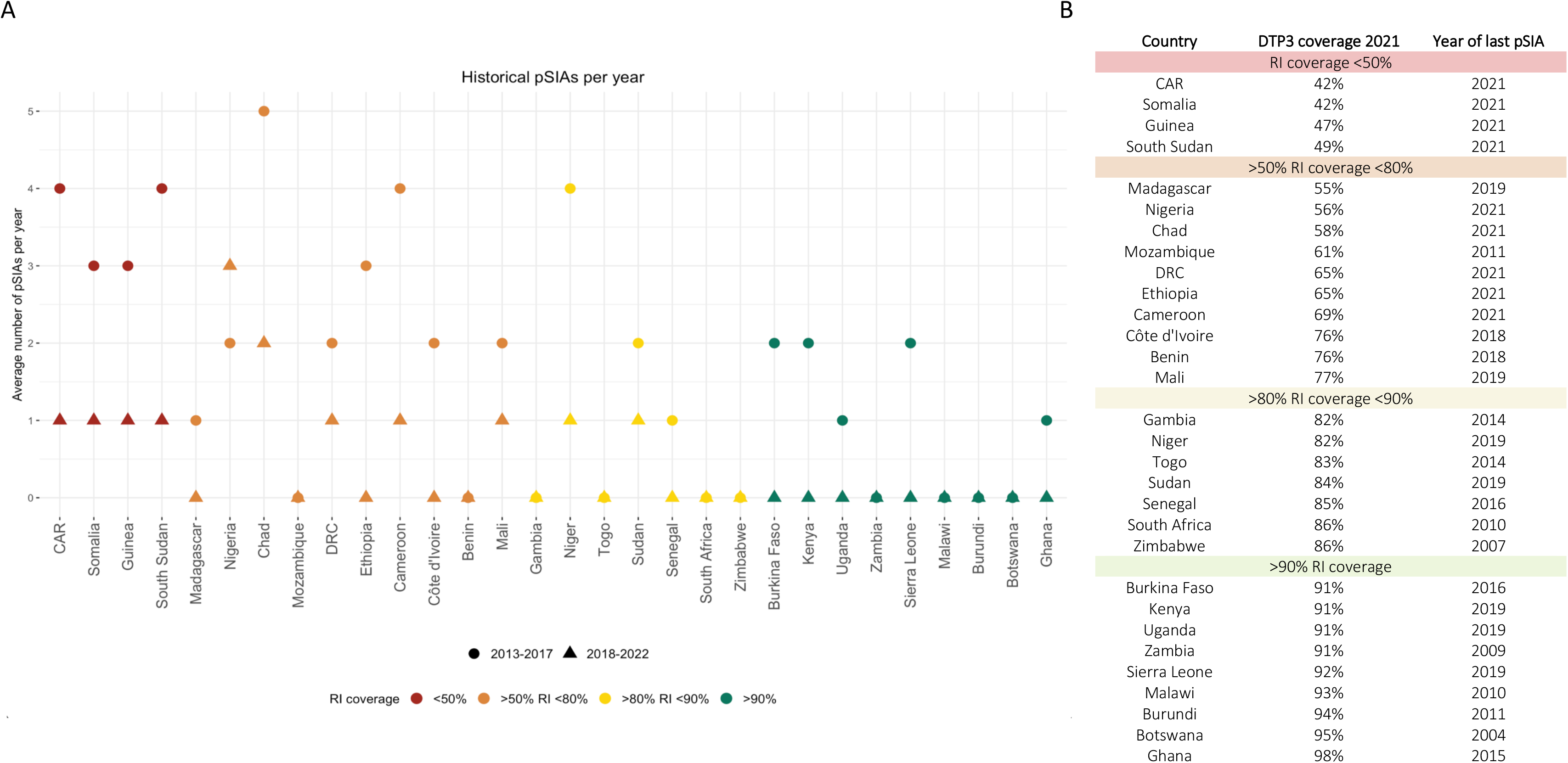
Historical pSIAs and RI coverage for 30 African countries. (A) Average number of pSIAs per year from 2013-2017 and 2018-2022 and (B) year of last pSIA and WUENIC estimates of DTP3 coverage. Preventative SIAs were defined as either a national or subnational immunisation day (NID, SNID) with bOPV (or tOPV pre-2016) and did not occur within 365 days of a WPV1 or VDPV1 detection, to distinguish historic pSIAs from oSIAs. Any SIAs that occurred within 365 days of a WPV1 or VDPV1 outbreak were not included in the pSIA count. CAR= Central African Republic; DRC = Democratic Republic of the Congo

Since the World Health Assembly’s 1988 resolution to eradicate polio by the year 2000, economic analyses have informed strategies to progress towards this goal. These studies inferred that polio eradication is cost-effective, assuming all vaccination stops following certification of global eradication (12–14). Together, polio vaccination and achieving global polio eradication are estimated to save approximately 40 million DALYs by 2050 (15). However, existing economic analyses do not consider the cost-effectiveness of specific operational decisions for preventative SIAs (pSIAs) and outbreak response SIAs (oSIAs) given a shrinking budget (16) or are outdated and apply to limited geographies (17). The distinction between pSIAs and oSIAs is important because pSIAs and oSIAs have different strategic goals and funding approaches, and programmatic decisions between vaccination strategies are required imminently, not just in the future context of OPV cessation. We aim to compare strategies of differing frequency of pSIAs to that of RI and oSIAs to identify at what levels of RI the risks of outbreaks and polio cases may outweigh the associated costs of implementing pSIAs. This is an evidence gap identified by stakeholders that is important to address if and when we approach the final few years of WPV1 transmission and start to consider OPV cessation.

## Methods

The GPEI works with a fixed annual budget for their operations, based on donations from stakeholders. This budget is used to support the GPEI’s objectives, including the global interruption of poliovirus transmission. Operationally, the planned budget is further divided into planned SIAs and outbreak response, and additional budget lines (not considered further in this study). Outbreak response includes oSIAs, and planned SIAs are either preventive in nature or used in endemic settings (not consider further here); pSIAs lower the risk of outbreaks due to the increase in population immunity. oSIAs include vaccination following a detected case (or at least two separate environmental detections that were collected from two different ES sites with no overlap in catchment area or collected from the same ES site at least two months apart (18)) and aim to raise population immunity in the affected areas to stop transmission. Operationally, pSIAs and oSIAs differ both in the target populations for vaccination as well as the funding and planning for activities – oSIAs must be implemented quickly and require more resource for rapid mobilisation. In this study, we consider only the risks of WPV1 outbreaks in a polio-free setting; the bOPV and IPV used in RI provide immunity against serotype 1. SIAs using bOPV also provide protection against infection as well as protection against paralysis. Outbreaks of vaccine-derived poliovirus serotype 2 require alternative vaccines and assumptions and are therefore beyond the scope of this analysis.

We evaluated different pSIA vaccination strategies in addition to RI and oSIAs for a population of 8 million children under five years of age, reflecting an average population size of a LMIC in Sub-Saharan Africa. We estimated costs alongside health benefits and risks for each vaccination strategy over a five-year time-period to align with the current GPEI timeline for polio eradication. Model outputs from each strategy include: probability of an outbreak, estimated cases of paralytic poliomyelitis and VAPP, number of outbreaks and estimated disability adjusted life years (DALYs) associated with these cases (Figure 2).

**Figure 2.**
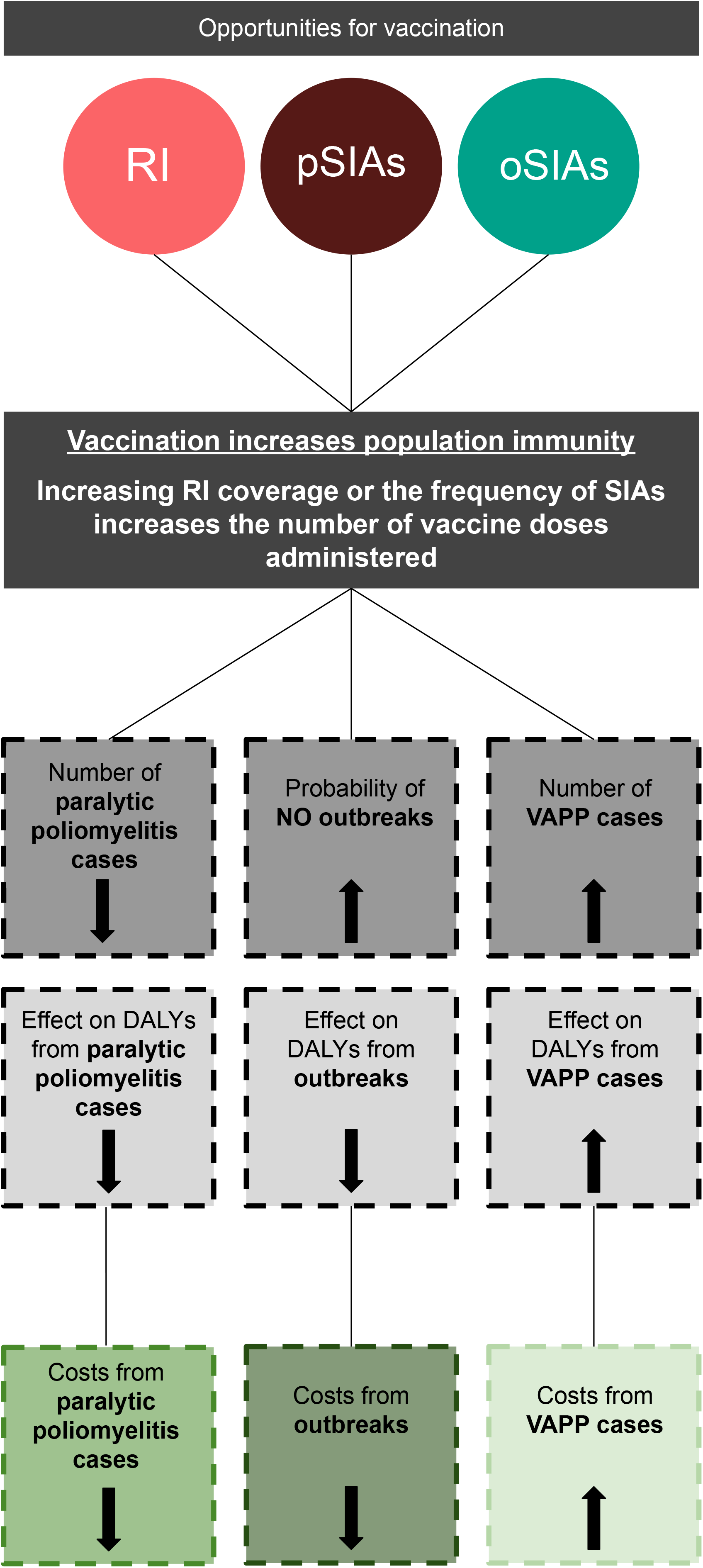
Health benefits, risks, and costs of polio vaccination. Schematic showing the relationships between vaccination, population immunity, paralytic poliomyelitis cases, adverse events and subsequent costs.

### Vaccination strategies

We explored three vaccination strategies (Table 1). We assume that vaccination via RI occurs in all strategies and follows a sequential immunisation schedule that includes 3 doses of bOPV given orally via drops in the mouth and 1 dose IPV administered intramuscularly or subcutaneously. The first two comparator strategies (RI+oSIAs+annual pSIAs and RI+oSIAs+biannual pSIAs) maintain the same assumptions about baseline RI coverage, but additionally include one pSIA either every year or every two years, respectively, across time.

**Table 1.** Polio vaccination strategies. RI = Routine Immunisation, oSIA = outbreak response supplementary immunisation activity, pSIA = preventative supplementary immunisation activity. An outbreak response was only conducted if a simulation had at least 1 case of paralytic polio.

| Vaccination strategy | Specification |
| --- | --- |
| Baseline strategy | RI + oSIAs<br>*No preventative SIAs* |
| Comparator strategy 1 | RI + oSIAs + *annual* pSIAs |
| Comparator strategy 2 | RI + oSIAs + *biannual* pSIAs |

In all strategies, we define an outbreak as at least one case of paralytic polio (18). An oSIA is conducted in all simulations for each strategy where at least one case was detected within any 90- day interval, in line with the median interval from notification to response as per the standard operating procedures (SOP) for polio outbreaks (18, 19). oSIAs continue until all cases are stopped over the five-year time horizon. We do not account for ES detections in this analysis. Children who were not vaccinated via RI, can receive additional doses of bOPV vaccine through either pSIAs or oSIAs. We assume that all SIAs reach 25% of children missed by RI, as prior evidence suggests that SIA coverage can be variable across locations and preliminary analysis of higher coverage for zero-dose children resulted in unrealistically high population immunity when compared to empirical data (20, 21). Appendix Table 4 includes a sensitivity analysis of different SIA assumptions.

### Model structure

We developed a stochastic non-linear mathematical model to simulate polio transmission dynamics, whereby infectious individuals develop either asymptomatic or symptomatic infection, both of which are assumed to be infectious. We specify vaccine induced immunity based on OPV and IPV doses (Appendix Figure 1). In the model, children under five years of age are either susceptible, fully vaccinated and protected from poliovirus infection, or have received an incomplete vaccination series (less than 3 bOPV doses + 1 IPV dose). Each subsequent dose of vaccine corresponds to additional protection and an opportunity for a child to seroconvert and be considered fully protected from poliovirus infection. Further explanation of model compartments and transitions is in Appendix Table 1.

### Time horizon and model assumptions

The modelled time horizon is five years, in line with the current GPEI strategic plan (22). The WPV1 case to infection ratio was assumed to be 1:200, which is consistent with estimates for poliovirus serotype 1 (23). For polio, the basic reproductive number, R_0,_ can vary substantially between locations depending on sanitation and hygiene conditions (24). Here we have used an R_0_ of 3 for the primary analysis, supported by recent work exploring the parameter space of variable R_0_ values in a non-endemic setting in Africa for children under five years of age (20, 25), and explore other R_0_ assumptions in a sensitivity analysis (Appendix Table 5). The proportion of children receiving three bOPV doses through RI at 6, 10, and 14 weeks is varied between 25%-100%, and for simplicity is assumed to occur upon entering the model. Children who have not seroconverted following vaccination or did not receive three or more doses of bOPV remain susceptible to infection. The proportion of children vaccinated with one dose of IPV is assumed to be equal to the third dose of bOPV RI coverage, which is in line with WUENIC data (26) (Appendix Figure 2). As of 2020, WHO recommends an additional IPV dose as part of RI (27), however, the number of countries that have introduced IPV2 as of 2023 are limited, therefore, we only consider IPV1 in our model.

We assume a fully mixed population of children under five years of age with no heterogeneity in the probability of a child being vaccinated in an SIA or in transmission of poliovirus. Simulations are run for 50 years before virus introduction, then one infection was introduced into the population at the start of the simulation and further importations were assumed to occur randomly at a rate of two importations per year. Different importation rates are reported in a sensitivity analysis in Appendix Table 6. The model was repeated for 1,000 stochastic simulations and run for five years. All models were run using the R package SimInf (28).

### Probability of an outbreak occurring

For each vaccination strategy, the probability of an outbreak was calculated using the proportion of stochastic simulations that resulted in at least one case of paralytic polio (i.e., a polio AFP case) over the entire modelled time horizon. The probability of an outbreak is comparable to maintaining elimination in an infection-free setting such as most African countries. This definition is not directly comparable towards the WHO criteria for elimination status (29), but is useful for understanding outbreak risk. We do not include the specific criteria for elimination status as modelling of the processes that lead towards achieving eradication are beyond the scope of this analysis.

### DALYs

DALYs were calculated for each strategy assuming that in LMICs, the average discounted lifetime DALYs associated with one paralytic poliomyelitis case, with no age-weighting is 14 DALYs per paralytic case of polio (30), assuming that one in 200 infections leads to irreversible paralysis and among those paralysed, 5-10% die when respiratory muscles become paralysed (31) and long-term mortality is approximately 20% higher in paralytic polio cases than the general population (32). The proportional contribution of YLD (years lost to disability) and YLL (years of life lost due to premature mortality) assuming 14 DALYs on average per case is 60% YLDs on average per case in addition to 40% YLLs on average per case (33). We assume no international transmission during outbreaks for calculations of DALYs.

### Health and economic outcomes

Incremental costs and incremental outcomes (in terms of DALYs and outbreaks averted) were estimated under each pSIA vaccination strategy in comparison to the baseline comparator (RI+oSIAs). We used a 3% discount rate for costs and 0% for health with no age-weighting, with other discounting assumptions explored in Appendix Figure 5. The total cost for each comparator strategy included baseline RI vaccine costs for both bOPV and IPV. We present both a health system (excludes GPEI costs), and GPEI perspective (includes only GPEI costs) for specification of costs, but do not include indirect costs of vaccination, such as opportunity costs of time spent for vaccination.

### Vaccine costs

Vaccine costs per dose for bOPV and IPV in Gavi supported countries were obtained from the latest UNICEF update in USD 2023, costing on average $0.18 and $2.00, respectively (34, 35) and administrative costs associated with polio RI were obtained from costs used in previous research (36), Appendix Table 3. All costs have been adjusted to 2023 USD. GPEI estimates OPV wastage ranges from 5-15% in SIAs and wastage is lower in SIAs than in RI (37). Estimates of IPV wastage vary between 5-20% in the literature (38) and country-specific analyses from Nigeria and The Gambia suggest IPV wastage can even be higher in practice, exceeding 20% for IPV wastage in some instances (39, 40). Accordingly, the main analysis assumes an average of 10% wastage for OPV in SIAs, 15% for OPV in RI and 15% for IPV, but further wastage assumptions are explored in a sensitivity analysis (Appendix Figure 6).

### Data and analysis on SIAs in Africa

The Polio Information System (POLIS) (41) was used to obtain SIA data for 2013-2022 and further analysis was done to distinguish preventative campaigns from outbreak response campaigns. We assumed that pSIAs included National or Subnational Immunisation Days (NID, SNID) with bOPV (or trivalent OPV (tOPV) pre-2016) that did not occur within 365 days of a WPV1 or VDPV1 detection by AFP surveillance or ES. Any SIA that occurred within this interval was not included in the overall count of pSIAs. Country level historic pSIA number and year of last pSIA were presented alongside WUENIC estimates of Diphtheria Tetanus Toxoid and Pertussis vaccine third dose (DTP3) coverage (42), an indicator of vaccination via RI as the DTP vaccine is administered concurrently with OPV in the routine immunisation series.

### SIA costs

The cost per child for both pSIAs and oSIAs was obtained from GPEI data (Appendix Table 7), for which an average across countries within the African Regional Office (AFRO) was used. Historic cost data for oSIAs has been less readily available, therefore, we assume oSIAs cost 2.0 times the cost of a pSIA, as per GPEI estimated costs for 2023. We also explore a range of proportional costs between pSIAs and oSIAs that align with previous assumptions in polio economic modelling (40) (Appendix Figures 7 and 8). Due to the stochasticity of epidemic simulations, there is considerable variability in outcomes for identical assumptions. The number of outbreaks, which affects total estimated costs, is variable and contributes to the variability in expected costs across all strategies (Appendix Figure 3).

### Adverse events

The expected risk of adverse events, such as vaccine associated paralytic polio (VAPP) in countries using OPV is 1 case of VAPP per 0.9 million doses of bOPV administered and declines with subsequent doses (43). VAPP rate across settings with and without SIAs has been shown to vary (44), although this variation is not well understood or documented. OPV administration following IPV receipt is associated with a further risk reduction of VAPP (43) and is accounted for in model assumptions (Appendix Table 3). VAPP cases contribute to overall risk and costs for each strategy. A VAPP case was considered equivalent to a case of wild-acquired paralytic polio for calculation of the expected costs of VAPP.

### Perspectives

Budget and incremental cost results are analysed from both the GPEI and health system perspectives. The GPEI costs include those associated with SIAs and IPV in RI. Treatment costs associated with paralytic polio (including VAPP) are paid for by the country and are included as health system costs. The probability of an outbreak under each vaccination strategy is analysed from both the global perspective for polio eradication and the country health system perspective.

### Ethics

Ethical approval for this project was received from the London School of Hygiene and Tropical Medicine, project ID 15873.

## Results

On average across all simulations, the expected number of WPV1 cases over five years is greatest in the baseline strategy (RI+oSIAs) and least in the annual pSIA strategy (RI+oSIAs+annual pSIAs) (Figure 3A). Under the base case assumptions (including R_0_ and proportion of zero dose children reached by SIAs) annual pSIAs achieve and maintain >80% probability of no outbreaks when baseline RI coverage is 50% (Figure 3B). The biannual pSIA strategy (RI+oSIAs+biannual pSIAs) achieves >80% probability of no outbreaks when RI is above 65% and the baseline strategy requires ≥75% RI coverage to achieve >80% probability of no outbreaks. The above results are from simulations assuming two importations per year during the five-year period of simulations. The frequency of importations was varied in a sensitivity analysis (Appendix Table 6); when RI was 50% or lower, the increased frequency of importation had a large increase on the probability of outbreaks, but for simulations with an RI ≥75%, the probability of no outbreaks remained above 80%.

**Figure 3.**
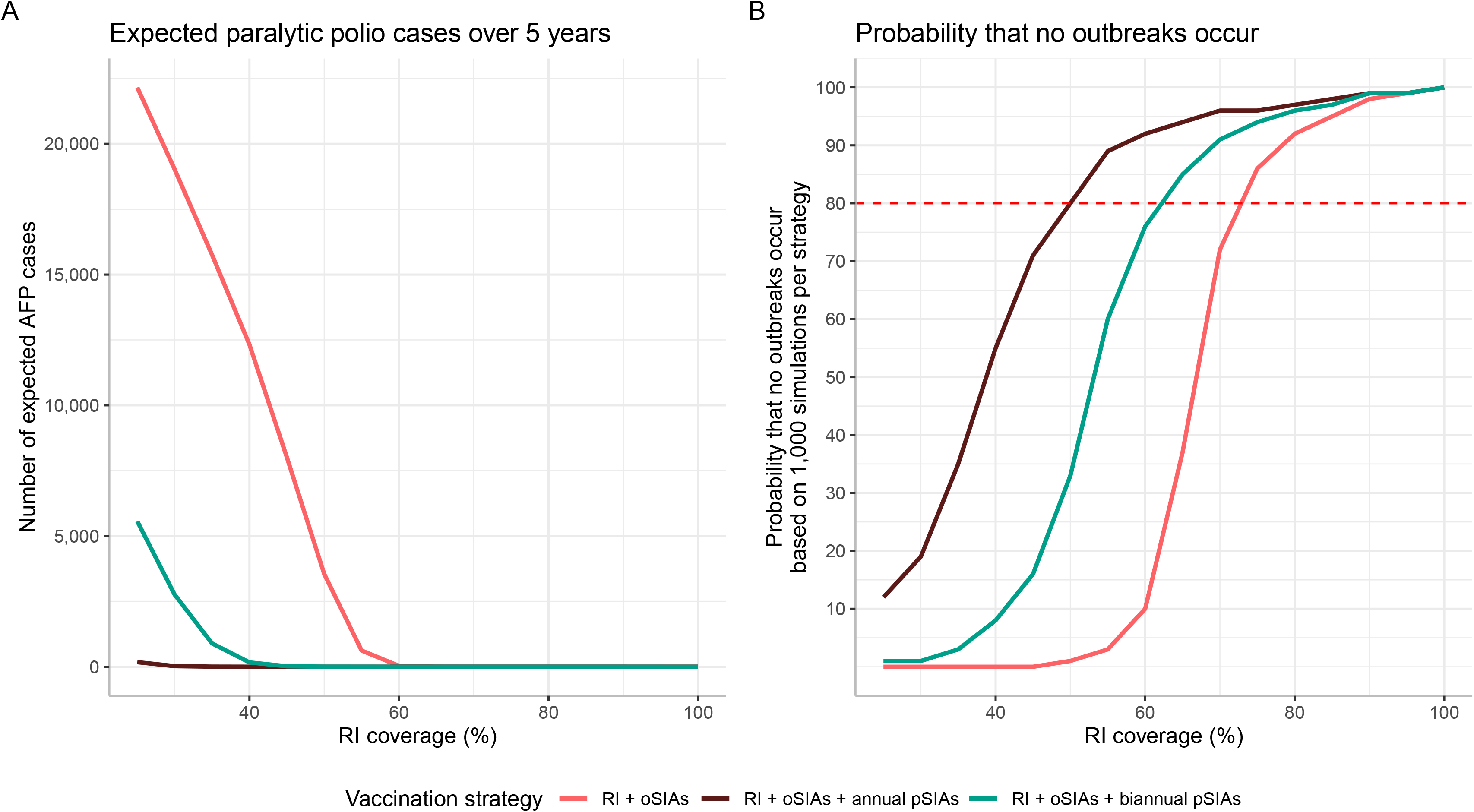
Estimated number of paralytic polio cases and probability that no outbreaks occur over the five-year modelled time horizon. (A) number of expected paralytic polio cases (presenting as a polio AFP case) and (B) probability of no outbreaks occurring across all vaccination strategies. Outbreak probability was based on 1,000 simulations per vaccination strategy. The red dashed line corresponds to 80% probability that no outbreaks occur.

Expected VAPP cases over five years are greatest in the annual pSIA strategy, corresponding to the strategy with the greatest number of vaccine doses administered, but fewest expected WPV cases over five-years. Estimated costs from both the health system and the GPEI perspectives are shown in Figure 4. From a health system perspective, estimated costs are lowest in the annual pSIA strategy due to fewer polio cases. From the GPEI perspective, estimated costs of the annual pSIA strategy always exceed the cost of the baseline strategy due to the number of preventative campaigns.

**Figure 4.**
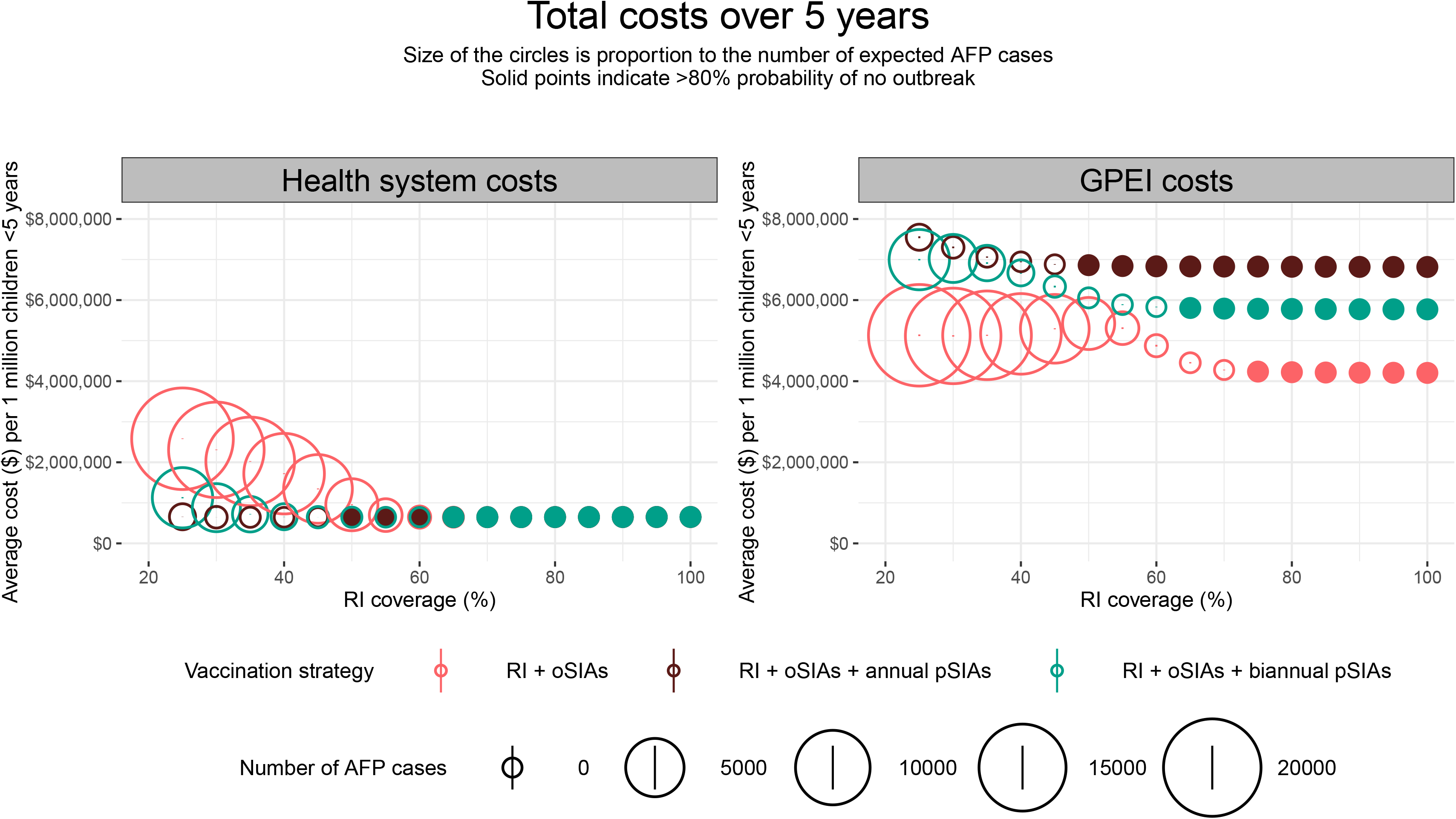
Health system and GPEI estimated total costs over five years. The size of the circle is proportional to the mean number of expected paralytic polio cases across all model simulations. The solid circles correspond to >80% probability the strategy had no outbreaks over a five-year period.

Incremental costs per DALY averted by both pSIA strategies in comparison to the baseline strategy are shown in Figure 5 with further explanation of the quadrants in a cost-effectiveness plane (Appendix Figure 9). For non-GPEI costs (Figure 5A), when RI coverage exceeds 50%, points for both pSIA strategies are densely clustered in the bottom-right quadrant highlighting approaches with relatively lower costs and more DALYs averted. Assuming herd immunity is calculated by (1 – 1/R_0_), when RI coverage exceeds 65%, the point when herd immunity is achieved in this simple homogenously mixed model, the annual pSIA strategy becomes less efficient in averting DALYS. For GPEI costs (Figure 5B), which do not account for treatment or RI bOPV costs, at all levels of RI coverage, the annual pSIA strategy incurs relatively higher costs in comparison to biannual pSIAs, with densely clustered points in top-right quadrant or along the positive y-axis. The cost effectiveness planes are presented in Appendix Figure 5 when a discount rate of 3% has been considered for both health and costs.

**Figure 5.**
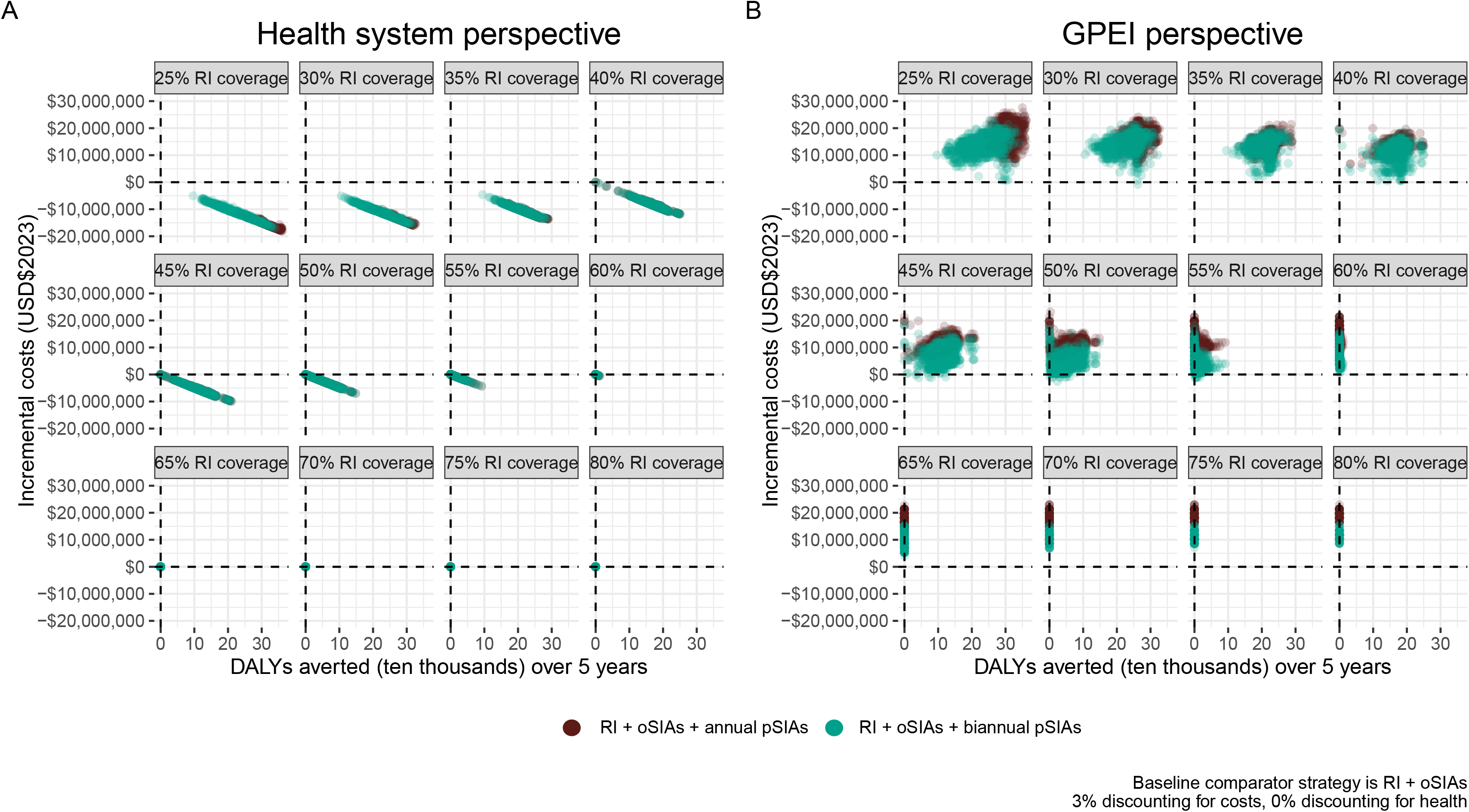
Incremental costs per DALY averted. Incremental costs and DALYs averted under the annual pSIA (RI+oSIAs+annual pSIAs) and biannual pSIA (RI+oSIAs+biannual pSIAs) strategies are compared to the baseline strategy (RI+oSIAs) assuming a 3% discount rate for costs and 0% discount rate for health. The points correspond to 1,000 model simulations. Here, it is assumed that the cost per child in an oSIA is two times the cost per child in a pSIA.

Incremental costs per outbreak averted capture the global perspective of polio eradication as a single outbreak under any vaccination strategy has implications for global polio eradication. This analysis has shown a greater number of outbreaks, albeit substantially smaller size, in the annual pSIA strategy in comparison to the baseline strategy when RI coverage is below 45% (under base-case assumptions), but the health system incremental costs are kept down by the small number of polio cases (Figure 6A). From the health system perspective, when RI coverage ranges from 45-70%, the annual pSIA strategy averts many outbreaks in comparison to the baseline strategy, but above 70% RI coverage, the annual pSIA strategy does not avert many outbreaks and is a costly approach. Both pSIA strategies avert few to no outbreaks when RI coverage exceeds 75%. Under the GPEI perspective, the annual and biannual pSIA strategies avert a similar number of outbreaks above 60% RI coverage, but annual pSIAs have higher incremental costs per outbreak averted, corresponding to the greater number of SIAs conducted (Figure 6B).

**Figure 6.**
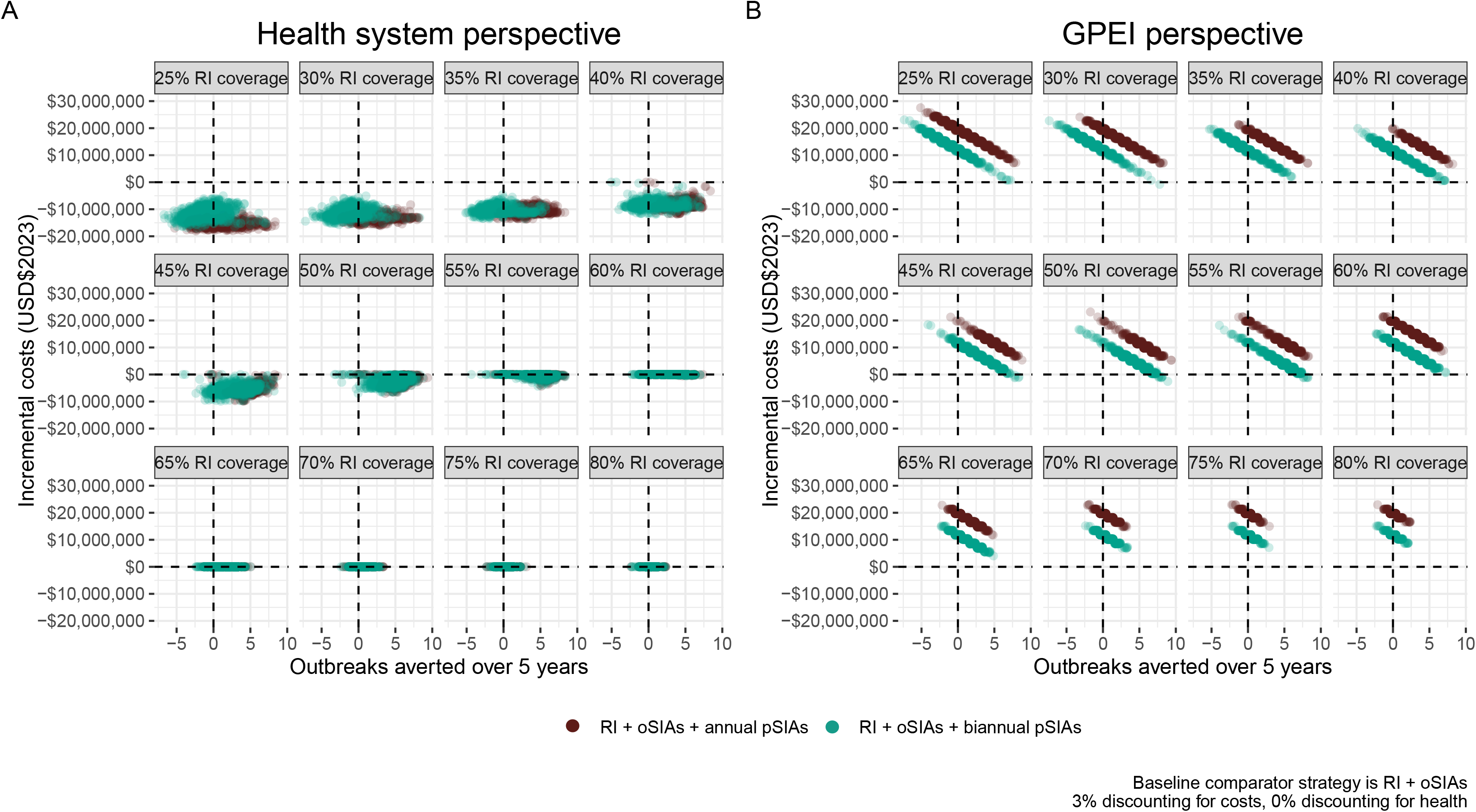
Incremental costs per outbreak averted. Incremental costs and outbreak averted under the annual pSIA (RI+oSIAs+annual pSIAs) and biannual pSIA (RI+oSIAs+biannual pSIAs) strategies in comparison to the baseline strategy (RI+oSIAs) assuming a 3% discount rate for costs and 0% discount rate for health. The points correspond to 1,000 model simulations. Here, it is assumed that the cost per child in an oSIA is two times the cost per child in a pSIA. An outbreak is defined as at least one case of paralytic polio. It is important to note that this figure does not suggest that the RI+oSIAs strategy is better performing at low RI coverage levels. Instead, negative (or no) outbreaks averted by both pSIA strategies at low levels of RI coverage correspond to two phenomena: (i) the RI+oSIA strategy has larger explosive outbreaks when RI coverage is low, so the susceptible population is depleted quicker, whilst the pSIA strategies have much smaller, albeit more frequent, outbreaks when RI coverage is low.

The implications for decision making are outlined in Table 2. Focussing on estimated risk if historic pSIAs have been regularly conducted, when RI coverage falls below 40%, the annual pSIA strategy averts many cases, so removal of pSIAs entirely would create substantial risk. However, pSIAs do not avert many outbreaks. It is important to note that this finding does not suggest that oSIAs are necessarily better performing at these coverage levels. Instead, the baseline strategy instead leads to quicker depletion of susceptibles or, results in more frequent vaccination via outbreak response following at least one paralytic case, whereas the pSIA strategies only vaccinate the target population annually or biannually. Countries with 50-90% RI coverage and historic pSIAs have a higher probability of no outbreaks occurring, with the outbreak risk reducing with higher RI. However, the risk of an outbreak is not removed entirely until the probability of no outbreaks reaches 100% (when WPV1 transmission is interrupted globally). All strategies require >95% coverage for 100% probability that no outbreaks occur, however, above 80% RI coverage, outbreak probability is low and, if an outbreak does occur, the expected outbreak size is very small. Figure 1 further summarises country-level historic pSIAs alongside the date since the last pSIA and current estimates of RI coverage. In summary, countries with <60% RI coverage have conducted more recent and frequent pSIAs and countries with >90% RI coverage have, on average, conducted fewer historic pSIAs per year. However, a few countries that fall below 80% RI coverage have not conducted a pSIA in over a decade.

**Table 2.** Policy implications of polio vaccination strategies. Implications for adopting each vaccination strategy based on a country’s RI coverage and historic SIA schedule according to GPEI SIA data (column 2). Average values across all model simulations are references for DALYs averted and outbreaks averted while expected paralytic polio cases are conditional averages amongst simulations that resulted in at least one case. The probability of no outbreaks occurring is obtained from the proportion of model simulations (out of 1,000 simulations) that resulted in zero paralytic cases and correspond to important implications for polio eradication.

| RI coverage | Did country conduct pSIAs in the past? | DALYs averted*? | Outbreaks averted*? | Estimated risk with annual pSIAs | Estimated risk with biannual pSIAs | Estimated risk relying on oSIAs only† | Implications for decision making |
| --- | --- | --- | --- | --- | --- | --- | --- |
|  |  |  |  | Average number of polio cases if an outbreak occurs‡<br>(Probability <u>no</u> outbreaks occur) |  |  |  |
| 35% | L | L | X | 5 (35%) | 894 (3%) | 15,753 (0%) | pSIA removal would have high risks and consequences |
| 50% | L | L | L | 1 (80%) | 4 (33%) | 3,549 (1%) | Removal of pSIAs altogether could lead to a high risk of outbreaks in subsequent years |
| 55% | L | L | L | 1 (89%) | 2 (60%) | 617 (3%) |  |
| 60% | L | L | L | 1 (92%) | 1 (76%) | 34 (11%) |  |
| 65% | L | L | L | 1 (94%) | 1 (85%) | 3 (37%) |  |
| 70% | L | X | L | 1 (96%) | 1 (91%) | 1 (72%) |  |
| 80% | L | X | L | 1 (97%) | 1 (96%) | 1 (92%) | Reducing the frequency of pSIAs could still maintain a low risk of large outbreaks |
| 90% | L | X | L | 1 (99%) | 1 (99%) | 1 (98%) |  |
| 100% | L | X | X | 0 (100%) | 0 (100%) | 0 (100%) | Even if pSIAs are removed, there is low to no risk of outbreaks |

\* Estimated DALYS and outbreaks averted are obtained from the annual pSIA strategy
† Estimated risk is obtained from the oSIA strategy assuming no pSIAs occur
‡ Expected paralytic polio cases were calculated by taking the average number of paralytic polio cases across all stochastic model simulations that had at least one case, so this is the average number of polio cases if an outbreak occurs

## Discussion

The key messages of our study are: (i) with higher RI coverage of three doses of bOPV and 1 dose IPV, the probability of outbreaks reduces considerably – under our model assumptions, outbreak size and risk are minimal when RI is above 65%. At lower values of RI, specific vaccination strategies should be considered; (ii) in scenarios where RI is below 50%, annual pSIAs limit outbreak risk and result in high cost, an annual pSIA strategy when RI is 50-80% results in a low probability of outbreaks and continues to be costly, (iii) biannual pSIAs require higher baseline RI coverage (65%) to achieve an equivalent probability of no outbreaks, but the reduction in cost has few negative outcomes; (iv) a strategy with only RI and oSIAs implicitly accepts some level of outbreak risk, but if RI is above 70-80% the risk of outbreaks is considerably less than other settings where RI is below this value.

At low levels (<50%) of RI coverage, the probability of outbreaks is high (99%) following an importation if pSIAs do not occur at any frequency. Further, the incremental costs show a clear benefit of a pSIA strategy, outweighing the increased number of expected VAPP cases and cost of vaccination. Countries such as Madagascar or South Sudan where 55% and 49% of the population under five years of age, respectively, are vaccinated with three or more DTP doses (used to approximate polio RI), have many subpopulations that could benefit from regular pSIAs. Indeed, between 2018-2022 South Sudan has implemented annual pSIAs, and Madagascar has implemented oSIAs in response to cVDPV1. Some countries in Africa have many subpopulations that fall towards the middle or upper range of analysed RI coverage estimates. In both Burkina Faso and Sierra Leone, for example, future SIAs would not seem necessary as routine immunisation coverage of three doses of bOPV and 1 dose IPV exceeds 90% without reliance on historic pSIAs, unless there are subpopulations with substantially lower coverage. The proportion of the population in Malawi that has received three doses of bOPV peaked in 2011, but has been unstable since, falling to 83% in 2016 (26), with no historic reliance on pSIAs. WPV1 was reported in Malawi and local transmission and international spread has since been reported. In our analysis, RI exceeding 80% would be unlikely to result in an outbreak, but we have not included heterogeneity in immunity in the simulations, likely over-estimating the probability of no outbreaks. It may also be that the frequency of WPV1 importation was under-estimated for the Malawi setting, and close monitoring of migration routes from endemic countries remains an important component of preventing polio outbreaks. Whilst WHO now recommends two doses of IPV (IPV2) as part of RI, many countries in Africa have not yet implemented this strategy. As the number of countries adopting this policy decision increases, future studies will be needed to analyse the impact of IPV2.

Our study has health policy implications for decision making on global polio eradication. Despite the high costs, the ability of pSIAs to decrease the probability of an outbreak is an important consideration for polio eradication. Should the GPEI adopt an annual pSIA strategy irrespective of estimated RI and importations? Annual pSIAs that include all children under five years of age in LMICs in Sub-Saharan Africa would consume most of the GPEI annual budget for activities and would be an inefficient use of funds, potentially reducing funds for other activities (surveillance, vaccination against other serotypes). However, prioritising SIAs in countries with low RI and perceived risk of introductions is a necessary compromise to which GPEI already adheres, and here we provide a framework to support decision making. When considering the vaccination strategies based on perceived outbreak risk (as described here) and the fixed budget for these activities, we must balance making the best use of limited resources while limiting WPV1 outbreak risk. The GPEI is reliant on a combination of country donations and philanthropic contributions to support activities, and budget has remained the same or reduced since 2010 and reduced further since the COVID-19 pandemic (45, 46). From the perspective of donors committing to polio eradication, reducing commitment prior to WPV1 interruption is a false saving: for every outbreak experienced, the timeline to WPV1 interruption become longer and the costs of eradication grows (22). Renewed commitment by donors was requested in 2022 (47) in light of the 2022-2026 Strategic Plan and these commitments remain essential to resource the activities needed to meet the objectives of polio eradication, including interrupting WPV transmission.

Our study has limitations. The main results assume a R_0_ of 3 in a homogenous population (both with respect to transmission and population immunity), a frequency of poliovirus introduction of two infections per year and SIAs reaching 25% of children missed by routine vaccination. These assumptions are a necessary simplification of real-world complexity. Several assumptions for R_0_, poliovirus introductions, and SIA reach were considered in a univariate sensitivity analysis. Should SIAs reach up to 50% of zero-dose children, the impact of SIAs on reducing outbreak risk is further reduced, consequently for the same costs a better outcome is achieved. Assuming a higher R_0_ and increasing the frequency of importations would also increase the outbreak risk. One of the most uncertain inputs of the analysis is importation rate: as poliovirus infection is typically asymptomatic this is not directly observable (although ES has frequently illustrated introductions in the absence of AFP cases (48)), and due to the changing epidemiology of polio globally, the importation rate will vary in time. We have not considered heterogeneity in the population. If there are pockets of the target population with higher rates of transmission and/or lower vaccination coverage, then the probability of an outbreak occurring would be higher. Additional discussion on the model framework is presented in Appendix Section 1, page 5.

Further, the model structure assumes that children are vaccinated when they are born into the population, not accounting for infection prior to vaccination. Because the average age of infection with poliomyelitis occurs after the vaccination schedule of bOPV given at 6, 10 and 14 weeks of age alongside the effect of maternal antibodies, this limitation is unlikely to impact the expected number of AFP cases. However, because the size of the birth cohort vaccinated may be overestimated in the absence of infant mortality assumptions, the costs of RI may be over-estimated here. Additionally, we do not include indirect costs of vaccination, such as opportunity costs of time spent for vaccination, which may result in an underestimation of costs across all strategies. The indirect benefits of these additional interventions (such as productivity increases from averting polio cases) are also not quantified in the costs and benefits of SIAs described here, therefore the deaths and burden averted are polio specific and exclude indirect economic effects, in contrast to some other analyses. Since 2020, pSIAs have been reduced or stopped altogether across most LMICs in Sub-Saharan Africa, meaning 2023 represents the fourth consecutive year of diminished immunity. This analysis does not directly assess the consequences of this diminished immunity but presents baseline research that can be used to inform these future research questions. We do not consider the costs of further delaying the eradication timeline through outbreaks, or the societal implications of outbreaks on polio eradication, both of which may further emphasise the need to implement pSIAs even when the outbreak risks are small. By limiting our analysis to a five-year time horizon, we underestimate the benefits of SIAs (particularly pSIAs) as they will increase the likelihood of eradication, meaning that control efforts after eradication can be scaled back (though not stopped). Further, the pSIA health system costs only consider the geographical remit stated in the model and ignores the potential for further international spread. International spread would be far more likely with larger outbreaks, consequently the health system costs are under-estimated.

In settings that have not recently experienced a WPV1 outbreak, it is challenging to decide between responding to short-term versus long-term risks, especially with a fixed budget. From the global perspective striving to achieve polio eradication, investing in pSIAs results in a greater probability of polio elimination, but still require justification in a pragmatic environment of finite resources. Therefore, future funding provisions and advocacy for the importance of pSIAs rely on research that communicates risk and burden in their absence. Here, we communicate the utility of different vaccination strategies and critically assess the risks associated with adopting different strategies given baseline RI coverage. Decisions made solely based on fixed budget, cost-effectiveness or burden reduction may not fully capture all consequences or benefits associated with adopting a particular vaccination strategy. Urgently, as importations of WPV remain a threat to the AFRO region, this analysis serves as an invaluable tool to estimate risk and plan vaccination activities across a range of settings.

## Data Availability

This analysis uses a transmission model to estimate all results presented in the manuscript. No metadata external to the model outputs are required. The code is documented on GitHub and is publicly available. All model parameters and assumptions used in the code are documented in the manuscript or appendix.(https://github.com/mauzenbergs/polio_econ) For computational efficiency, the repository also includes cleaned model outputs. To validate our results, those interested can use the data files in the repository and the accompanying processing code to validate economic results.

## Authors’ contributions

MA and KMOR conceptualised the study. MA curated the data, conducted the modelling analysis, prepared visualisations of results and wrote the original draft. MA, KMOR and KA independently validated the data and results. All authors contributed to reviewing results and editing of the manuscript and have approved the final version. The authors alone are responsible for the views expressed in this article and they do not necessarily present the decisions, policy or views of their affiliated organisations.

## Acknowledgements

This project was funded by the Bill and Melinda Gates Foundation (BMGF), OPP1191821. We would like to thank Rachel M. Burke (BMGF), Grace Macklin (WHO), John Edmunds (LSHTM), Anna Vassall (LSHTM), Stephan Widgren (SimInf), and others in the GPEI Subgroup for Analysis and Modelling for their time and feedback over the course of this project.

## Data sharing

Simulation code used in the analysis and sample model outputs can be accessed at: https://github.com/mauzenbergs/polio_econ.

## Acronyms

Supplementary Immunisation Activities (SIAs), outbreak response SIAs (oSIAs), planned preventative SIAs (pSIAs), Routine Immunisation (RI), Global Polio Eradication Initiative (GPEI), bOPV (bivalent oral polio vaccine), IPV (inactivated polio vaccine), circulating vaccine derived poliovirus (cVDPV), vaccine associated paralytic polio (VAPP), disability adjusted life years (DALYs), wild poliovirus serotype 1 (WPV1), WHO (World Health Organization), The United Nations Children’s Fund (UNICEF), WUENIC (WHO and UNICEF estimates of national immunization coverage), acute flaccid paralysis (AFP), environmental surveillance (ES), African Regional Office (AFRO), Bill and Melinda Gates Foundation (BMGF), low- or middle-income country (LMIC), London School of Hygiene and Tropical Medicine (LSHTM), standard operating procedures (SOP).

