## Appendix for "Outbreak risks, cases, and costs of vaccination strategies against wild poliomyelitis in polio-free settings: a modelling study"

#### Table of Contents

Section 1: Model structure, parameters and limitations

i. *Appendix Figure 1 – SIR model structure*
ii. *Appendix Table 1 - Model compartments and transitions*

Section 2: Model parameters and cost assumptions

i. *Appendix Figure 2 – WUENIC coverage estimates supporting vaccination coverage assumptions*
ii. *Appendix Table 2 – Model parameters*
iii. *Appendix Table 3 – Cost inputs*

Section 3: Stochastic simulations result in variable outcomes across strategies

i. *Appendix Figure 1 – Variable number of outbreaks across strategies*
ii.
iii. *Appendix Figure – Average number of expected AFP cases, expected VAPP cases and DALYS across all vaccination strategies*

Section 4: Sensitivity analyses

i. *Appendix Figure 5 – Sensitivity analysis: 3% discounting for health*
ii. *Appendix Figure 6 – Sensitivity analysis: budget impact under different wastage assumptions*
iii. *Appendix Figure 7 – Sensitivity analysis: incremental costs and DALYs averted under different oSIA cost assumptions*
iv. *Appendix Figure 8 – Sensitivity analysis: incremental costs and outbreaks averted under different oSIA cost assumptions*
v. *Appendix Table 4 – Sensitivity analysis: model outputs under different assumptions about the proportion of children reached by SIAs*
vi. *Appendix Table 5 – Sensitivity analysis: model outputs under different assumptions about $R_0$*
vii. *Appendix Table 6—Sensitivity analysis: model outputs under different assumptions about importation rate*

Section 5: Supporting information

i. *Appendix Table 7—Proportional costs between pSIAs and oSIAs*
ii. *Appendix Figure 9 – Cost-effectiveness quadrants for interpretation of incremental results*

### Section 1: Model structure, parameters and limitations

#### Model structure

A stochastic non-linear mathematical model was used to simulate polio transmission dynamics, whereby infectious individuals develop either asymptomatic (I, or infectious, compartment in Appendix Figure 2), or symptomatic infection (C, or case, compartment), both of which are assumed to be infectious. Both infections and cases recover to the Rn compartment. In the model, children are either susceptible (S0 compartment), fully vaccinated and protected from poliovirus infection (Rv compartment) or have received an incomplete vaccination series (less than 3 bOPV doses + 1 IPV dose, modelled separately). Children who were not vaccinated via RI, can receive additional doses of bOPV vaccine through either pSIAs or oSIAs. Each subsequent dose of vaccine corresponds additional protection and an opportunity for a child to seroconvert and be considered fully protected from poliovirus infection (corresponding to each of the Sn tiers).

The left side of Appendix Figure 1 corresponds to children missed by RI who have an opportunity for additional bOPV doses via SIAs. Progression through the model to different Sn compartments happens only at the time of an SIA.

The right side of Appendix Figure 1 corresponds to RI and happens daily, when life births enter the population. Children vaccinated via RI are assumed to receive a sequential schedule of both bOPV and IPV, whereby they are assumed to seroconvert and be protected from poliovirus (transitioning to Rv compartment). Only children previously vaccinated via RI are eligible for vaccination with IPV and IPV is provided alongside the third dose of bOPV via RI in most African countries.

In the main analysis, we assume that IPV = bOPV coverage, but if IPV<OPV, children who were vaccinated with bOPV doses via RI, but did not complete their vaccination series and receive IPV, could be eligible for additional bOPV doses via SIAs, representing compartments in the grey shaded box.

To guide understanding of the model diagram, the compartments correspond to the following:

- S0 = susceptible to poliovirus, no vaccine received
- I = infectious, but asymptomatic
- C = infectious and polio (AFP) case
- Rn = recovery via natural infection
- $S_n$ = susceptible to poliovirus infection, but have received n bOPV doses via SIAs
- $RI_{3bOPV + IPV}$ = vaccinated with 3 bOPV doses + IPV at birth via RI
- $S3_{RI}$ = vaccinated with bOPV, but missed IPV dose

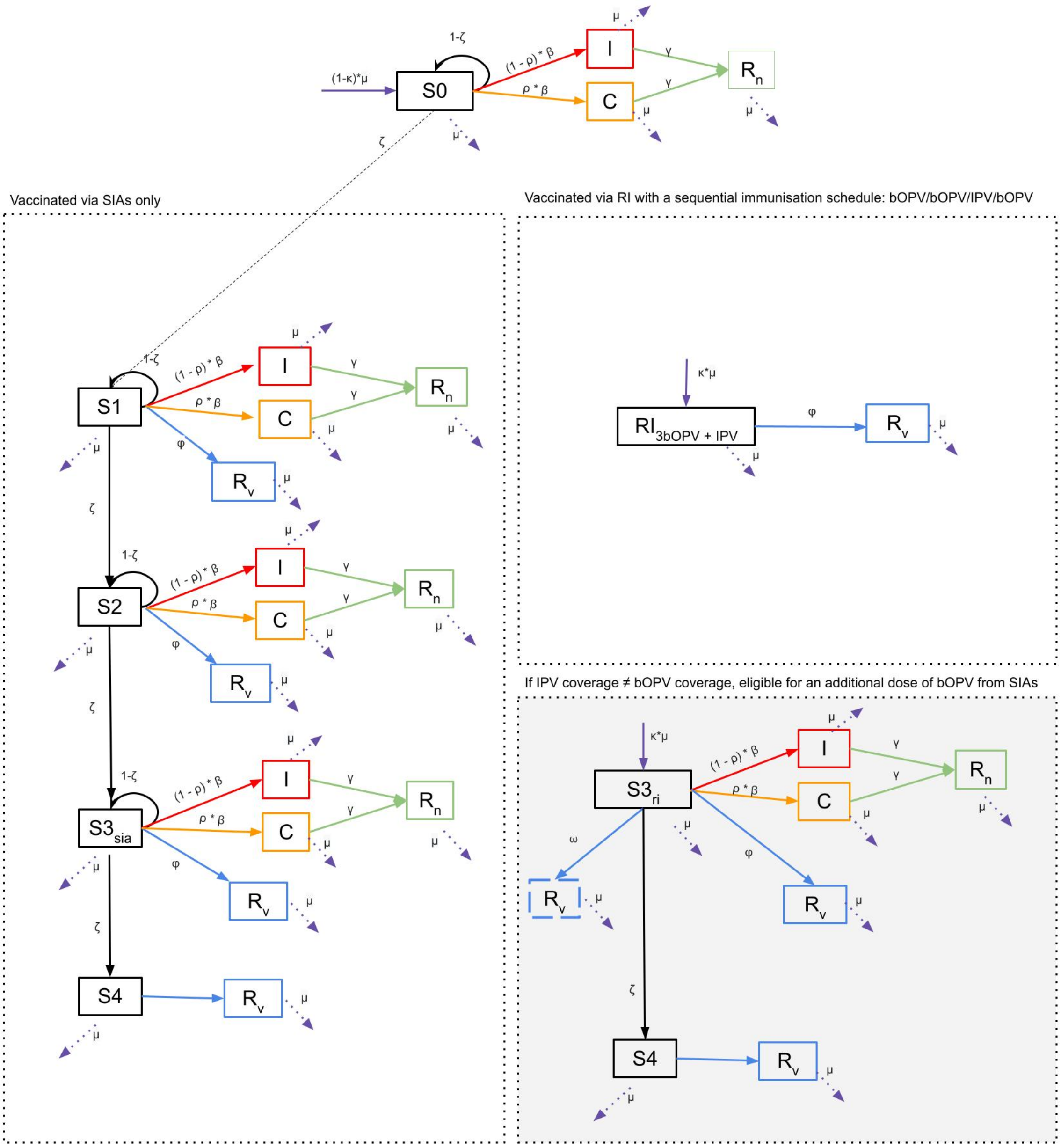


*Appendix Figure 1. Model structure of the extended SIR model. Births enter the population either via the S0 compartment or the $S3_{ri}$ compartment, assuming vaccination with three bOPV doses occurs via RI at the time of birth. Children can exit the population from any compartment via death. Children missed by RI at birth remain in the S0 compartment, with potential for vaccination with one dose of bOPV via pSIA or oSIA. Following vaccination during an SIA, a child can then progress to the S1 compartment. Each of the SIA tiers (S1, S2, S3, S4), correspond to the number of bOPV doses received via an SIA and allow for potential seroconversion and transition to the Rv compartment according to assumed bOPV vaccine efficacy at any one of these tiers. Children who do not seroconvert following bOPV during an SIA progress to the next vaccination tier and await subsequent SIA vaccination. Each of the SIA tiers remain susceptible to infection. In this model we assume that bOPV coverage = IPV coverage, however, the model is set up such that if these coverage values are not equal, if a child receives three bOPV doses but no IPV dose, they remain eligible for a 4th bOPV dose via SIAs (grey shaded box).*

#### Model compartments and transitions

A review of the immunogenicity of vaccination schedules with both IPV and three doses of bOPV indicate high rates of seroconversion alongside some geographical variability [1] and sustained protection against paralytic poliomyelitis [2]. Accordingly, we assume seroconversion and transition to the Rv compartment is 100% for children who received three doses of bOPV and one dose of IPV and no waning of protection. The assumptions described here about vaccination activities provide a 'steady state' immunity profile which we use to explore the effects of polio introductions. We assume that once five years of age is reached, children are no longer relevant to poliovirus transmission, due to the high likelihood of immunity against infection due to vaccination.

*Appendix Table 1. Model compartments and corresponding explanation of compartmental transitions and assumptions*

| Compartment | Assumption |
|---|---|
| S0 | Individuals in this compartment have not been vaccinated, i.e., zero doses received of both bOPV and IPV. The proportion of unvaccinated individuals born into this compartment depends on assumed RI coverage. From S0, individuals exposed to 1 dose of bOPV via an SIA progress to S1. Individuals who stay in S0 remain susceptible to infection. |
| S1 | Individuals in this compartment have received 1 dose of bOPV via an SIA. Individuals in S1 did not receive 3 doses of bOPV via RI at birth, therefore, their only opportunity for vaccination was via an SIA. From S1, a certain proportion will seroconvert after 1 dose and transition to Rv, according to bOPV vaccine efficacy (see parameter $\phi$ below). While those that did not seroconvert stay in S1 and remain susceptible to infection or able to be vaccinated in a subsequent SIA. |
| S2 | Individuals in this compartment have received 2 doses of bOPV via an SIA. From S2, a certain proportion will seroconvert after 2 doses and transition to Rv, according to bOPV vaccine efficacy (see parameter $\phi$ below). While those that did not seroconvert stay in S2 and remain susceptible to infection or able to be vaccinated in a subsequent SIA. |
| $S3_{sia}$ | Individuals in this compartment have received 3 doses of bOPV via an SIA. From $S3_{sia}$, a certain proportion will seroconvert after 3 doses and transition to Rv, according to bOPV vaccine efficacy (see parameter $\phi$ below). While those that did not seroconvert stay in $S3_{sia}$ and remain susceptible to infection or able to be vaccinated in a subsequent SIA. |
| S4 | Individuals in this compartment have received 4 doses of bOPV and therefore, 100% are assumed to seroconvert and transition to Rv. |
| $RI_{3bOPV + IPV}$ | Assuming that bOPV coverage = IPV coverage. Children in this compartment are vaccinated via RI with a sequential schedule of OPV/OPV/IPV/OPV. It is assumed after 3 doses of bOPV and 1 dose IPV 100% of children seroconvert and transition to the Rv compartment and are also protected from paralysis. |
| $S3_{ri}$ | Individuals in this compartment received 3 doses of bOPV at birth via RI. From $S3_{ri}$, a certain proportion will seroconvert after 3 doses and transition to Rv, according to bOPV vaccine efficacy (see parameter $\phi$ below). If IPV coverage ≠ bOPV coverage, those who did not seroconvert after 3 doses of bOPV are eligible for vaccination with 1 dose of IPV, depending on assumed IPV coverage (see $\omega$ parameter below), or receive a 4$^{th}$ bOPV dose from an SIA. Of those vaccinated with IPV, 100% are assumed to seroconvert and transition to Rv |
| I | Individuals in this compartment are infectious, but asymptomatic. They can spread infection to others but are not a paralytic case of polio. Importations of infection who enter the population enter via this compartment. |

| | |
|---|---|
| C | Individuals in this compartment are a paralytic case of polio. They can spread infection to others and trigger an outbreak response campaign in the RI+oSIA strategy. |
| Rn | Individuals in this compartment have recovered from natural infection and are assumed immune to subsequent infection. |
| Rv | Individuals in this compartment have received sufficient vaccination to result in seroconversion and are assumed immune to subsequent infection. |

#### Model framework

It is important to discuss the model framework used in this analysis and associated pros and cons of the simplistic model structure and assumptions. Model simplicity allows for easy interpretation and is applicable to a range of settings. Here, we model a hypothetical population size for a LMIC in sub-Saharan Africa using certain fixed parameters, but the simple model structure can be easily adapted to fit specific countries, or even subnational populations. The model compartments and transitions are easy to understand by a wide range of audiences, not just mathematical modellers and the SimInf package allows for easy adaptation of scheduled vaccination activities to local contexts. The SimInf package also does not require extensive coding experience of complex transmission models and differential equations and the available model code associated with this analysis is easily reproducible. To reach a wide audience of stakeholders involved in global polio eradication, the simplicity of model structure was an important consideration for this analysis.

The simplicity of the model does give rise to several limitations. In this model, we assume homogenous mixing, assume that the same polio programme has been in place for 50 years prior to model initiation, assume SIAs reach 25% of children unvaccinated by RI, assume a single value for $R_0$ and other model parameters with no uncertainty. Of particularly importance is heterogeneity in the population structure. If there are pockets with higher rates of transmission or lower vaccination coverage (or both) then this would increase the likelihood of outbreaks and thus decrease the likelihood that eradication will be achieved. Sensitivity analyses in Appendix Section 4 further explore varying assumptions about SIA target population, importation rate and $R_0$. In summary, under different assumptions, model outputs across vaccination strategies are only affected when RI coverage is very low and therefore, support the assumptions used in the main analysis.

### Section 2: Model parameters and cost assumptions

#### IPV and OPV coverage estimates

This WEUNIC data shows the relationship between IPV and OPV3 coverage (three doses of bOPV vaccine) for twenty-five countries in sub-Saharan Africa. IPV was introduced later in time in all countries, and although at the time of introduction IPV coverage was lower than OPV3 coverage in most countries, in 2021, IPV and OPV3 coverage was roughly equal across all countries. This data supports the model assumption that IPV coverage = OPV3 coverage, or all children that receive a third dose of bOPV, also received IPV via RI.

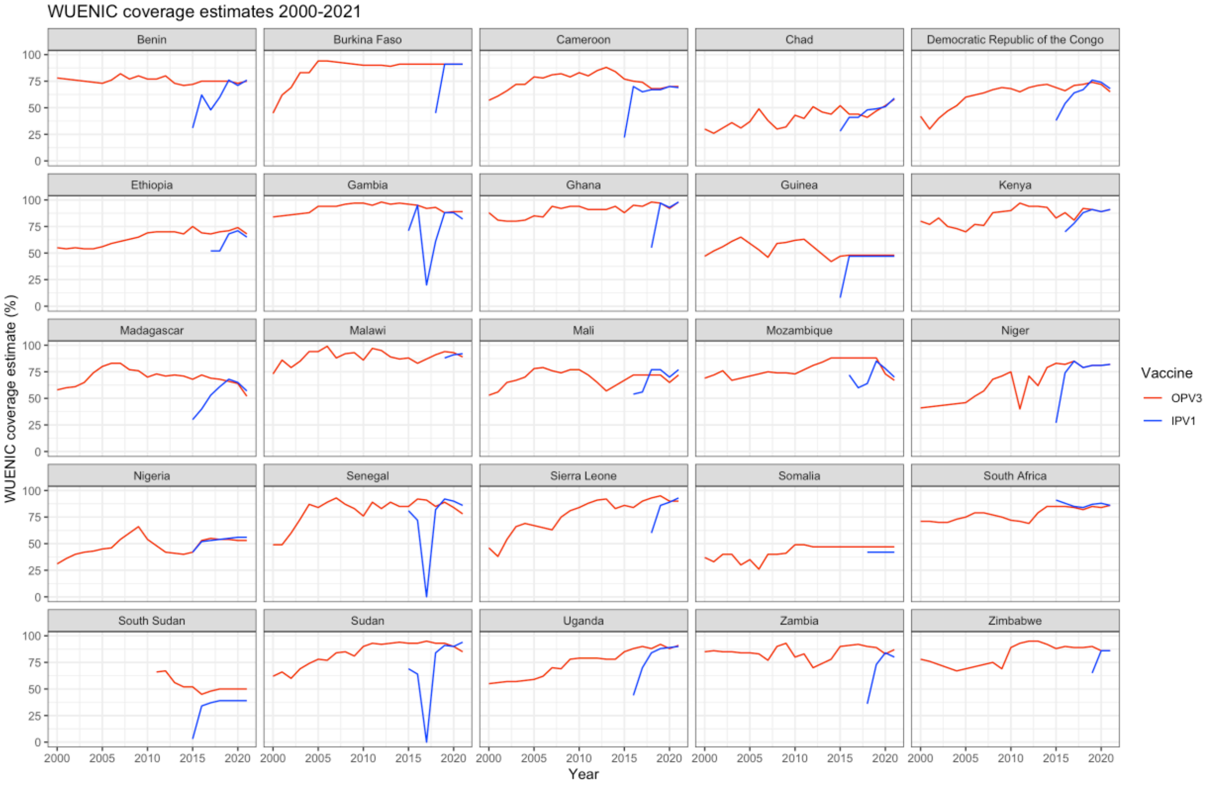


*Appendix Figure 2. WUENIC coverage estimates 2000-2021 for OPV3 and IPV1 across twenty-five low-and-middle income Sub-Saharan African countries. The strong correlation between OPV3 and IPV1 coverage estimates in 2020 and 2021 (when scale-up of IPV implementation was reached) support model assumptions that OPV coverage is equal to IPV coverage.*

#### Model parameters and assumptions

*Appendix Table 2. Table of model parameters and corresponding values and assumptions*

| Parameter | Value | Assumption |
|---|---|---|
| γ | 1/8 | γ = recovery rate – the infectious period for polio is between 7-10 days before AND after symptoms. Here, we took the average, 8 days that an individual can remain in the I or C compartment, before transitioning to $R_n$ |
| $R_o$ | 3 | Basic reproductive number |
| β | 0.375 | Calculated knowing R0 is 3, and recovery rate is 1/8 |
| ρ | 1/200 | Case to infection ratio for WPV |
| κ | Varied between 0.25 – 1.0 | Proportion that received 3 doses of bOPV via RI |
| ζ | 0.25 | Proportion of the target population reached by the SIA, those missed by the campaign are represented by 1- ζ |
| ω | Varied between 0.25 – 1.0 | Proportion exposed to IPV vaccination |
| ϕ | $1 - ((1 - 0.5)^{doses})$ | Vaccine effectiveness of bOPV for protection against serotype 1, i.e., the proportion of the population that seroconvert and transition to Rv assuming vaccine efficacy (VE) is 50% |
| μ | $5*10^{-4}$ * population | Birth rate = death rate<br>4,000 live births per day in a country of 8 million is the average number of live births across African countries |
| Herd immunity | $1\text{-}1/R_0$ | Using an $R_0$=3, the herd immunity threshold would be assumed to be reached at 66.67% RI coverage |

#### Cost assumptions

We assume costs of SIAs are the same across the entire modelled time horizon. Costs associated with vaccine doses and the number of children vaccinated during an SIA are estimated using the entire target population of eight million children under five years of age, even though in practice, the true proportion of children reached during SIAs is often much lower due to operational challenges and other logistical shortcomings [3]. As model outputs captured the variability across all stochastic simulations, costs for each individual simulation were calculated across all strategies. The average expected costs for each strategy were then obtained by taking the mean cost across all simulations for each corresponding strategy. The GPEI costs include those associated with SIAs and IPV in RI. Treatment costs associated with paralytic polio (including VAPP) are paid for by the country and includes as health care system costs.; specific equations used in the costing estimates can be found below in Appendix Table 3.

Literature has shown that costs associated with RI administration can vary across RI coverage for different antigens [4, 5], but usually the threshold cut-off is below and above 80% RI coverage and explicit evidence of this differential for polio is not well documented. Accordingly, we use one value for costs associated with RI administration for polio as documented by Kalkowska et al.

*Appendix Table 3. Table of cost inputs and corresponding values and assumptions*

| Item | Value (USD$2023) | Assumption / reference |
|---|---|---|
| bOPV dose | $0.18 | Average cost for a GAVI country [5], price per dose in a 10-dose vile |
| IPV dose | $2.00 | For a GAVI country 10-dose vile and translated into USD$2023 [6] |
| RI OPV admin | $1.06*3 doses | Costs associated with administration, procurement and storage of OPV for use in RI. 2019 costs were obtained from appendix Table A1 [7]. We assume costs and wastage are based on the size of the entire birth cohort. |
| RI IPV admin | $2.00 | Costs associated with administration, procurement and storage of IPV for use in RI. 2019 costs were obtained from appendix Table A1 [7]. We assume costs and wastage are based on the size of the entire birth cohort. |
| pSIA | $0.50/child | Includes operational costs, social mobilisation and administration. We assume that SIAs are planned to reach 100% of the population and costed accordingly but the actual population vaccinated is 25% due to operational challenges. Costs are obtained from GPEI |
| oSIA | $1.0/child<br><br>Range: $0.50 – $1.50/child | Includes costs associated with emergency response to an outbreak. We assume that SIAs are planned to reach 100% of the population and costed accordingly but the actual population vaccinated is 25% due to operational challenges. Costs are estimated based on GPEI costs for pSIAs |
| VAPP rate | Dependent on dose and vaccination schedule | 1st dose: 0.9 VAPP cases/1 million bOPV doses administered, a 6.6-fold greater risk than following subsequent doses [8]<br><br>The risk of VAPP following OPV vaccination is reduced if IPV has been received. We assume no risk of VAPP associated with IPV vaccination and a 53% reduction in |

| | | |
|---|---|---|
| | | VAPP cases following bOPV if IPV has already been received |
| DALY | 14/paralytic case | Assume AFP case = VAPP case |
| Paralytic case | $700/case | Assume AFP case = VAPP case, cost is for a low-income country [4] |
| bOPV wastage RI | Range 10-20% | Range of estimated wastage of bOPV in RI [9, 10] |
| bOPV wastage SIA | Range 5-15% | Range of estimated wastage of bOPV in both pSIAs and oSIAs [9, 10] |
| IPV wastage RI | Range 5-20% | Range of estimated wastage of IPV in routine settings. Range varies based on vial size and setting [9, 10] |
| Health care system & non-GPEI costs | | Treatment costs (red) + RI OPV costs (blue)<br><br>Cost per AFP case + Cost per VAPP case<br>+<br>(Births * OPV wastage for RI * Cost per dose of bOPV * total doses received per child * RI costs for OPV administration) |
| GPEI costs | | SIA costs (green) + RI IPV costs (orange)<br><br>(SIA target population * OPV wastage for SIAs * Cost per dose of bOPV * Number of SIAs * SIA cost per child)<br>+<br>(Births * IPV wastage for RI * Cost per dose of IPV * RI costs for IPV administration) |

#### Section 3: Stochastic simulations result in variable outcomes across strategies

##### Variable number of outbreaks across all strategies

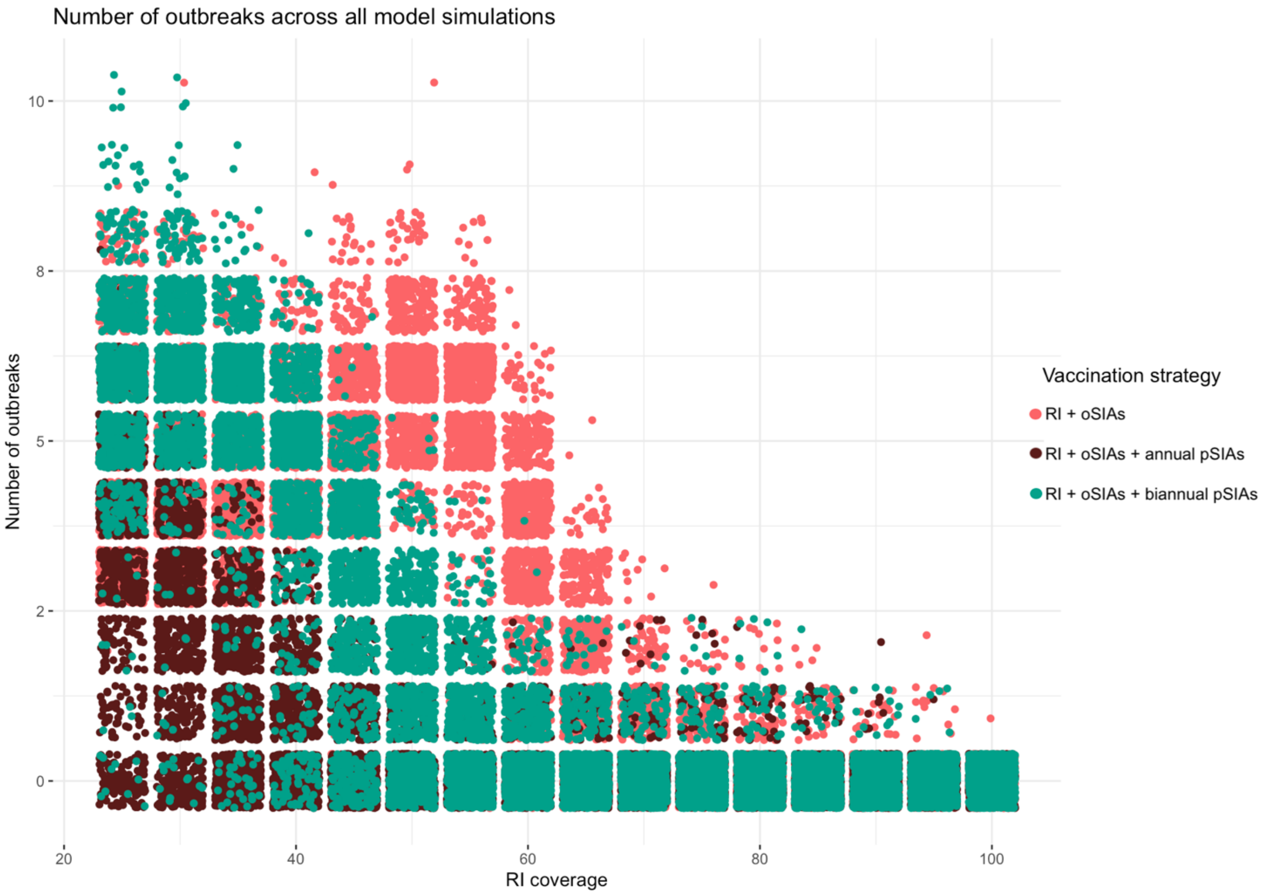


*Appendix Figure 1. Number of outbreaks in each individual model simulation for each vaccination strategy, among simulations with ≥1 paralytic polio case. The low number of outbreaks at RI coverage levels >70% correspond to a lower probability of an outbreak, which was lowest in the annual pSIA strategy.*

The stochasticity of epidemics means that there is considerable variability in outcome for identical assumptions focussing on simulations that had ≥1 paralytic polio case (Appendix Figure 1). We define an outbreak as at least one case of paralytic poliomyelitis, which we assume would present as acute flaccid paralysis (AFP). To count the total number of outbreaks, all cases that occurred within 90 days of the first case were considered part of the same outbreak. Cases that occurred in subsequent intervals of 90 days were considered part of different independent outbreaks. Even though some cases that are allocated to different outbreaks may in fact be linked or a part of the same outbreak, the 90-day threshold aligned with the schedule of oSIAs and was used as a consistent unit to count outbreaks across strategies. This method of counting outbreaks may incorrectly attribute linked cases from the same outbreak as independent outbreaks but was taken to reduce model complexity. As standard operating procedures for polio outbreaks recommend an oSIA in affected areas within 90-days of the first case, if an outbreak is not stopped or subsequent cases arise later, this would trigger a subsequent oSIA, a chronology which is accurately accounted for in this analysis. In the absence of having uniquely identifiable outbreaks within the model, simplifying the outbreak count by using a 90-day cut-off is a limitation that may over-estimate the total number of outbreaks, however, the counting methodology is consistent across all evaluated strategies.

#### Trends in VAPP and DALYs

Expected VAPP cases over the same interval are greatest in the annual pSIA strategy, corresponding to the strategy with the greatest number of vaccine doses administered (Appendix Figure 4). The annual pSIA strategy administers the greatest number of vaccine doses and is the strategy that results in the fewest expected WPV cases over five-years (main text figure 2A). At RI coverage levels below 70%, DALYs incurred are greatest in the RI+oSIA strategy, the strategy with the greatest number of expected AFP cases. However, at RI coverage levels >70%, the average number of VAPP cases exceeds the number of WPV1 cases in all pSIA strategies, which drives these two strategies to have a greater number of estimated DALYs lost than the RI strategy alone. Expected DALYs in the annual pSIA strategy are greatest when RI coverage exceeds 70% because the number of pSIAs remains constant over time, resulting in more VAPP cases and consequently, a greater number of DALYs incurred. In contrast, above 70% RI coverage, the RI+oSIA strategy has fewer WPV1 cases and no or fewer oSIAs, resulting in fewer VAPP cases and therefore fewer expected DALYs incurred. When more optimistic assumptions about SIA coverage were made, results followed similar trends and the only substantial difference between expected AFP cases and outbreak probability was at RI coverage levels below 35% (Appendix Table 4).

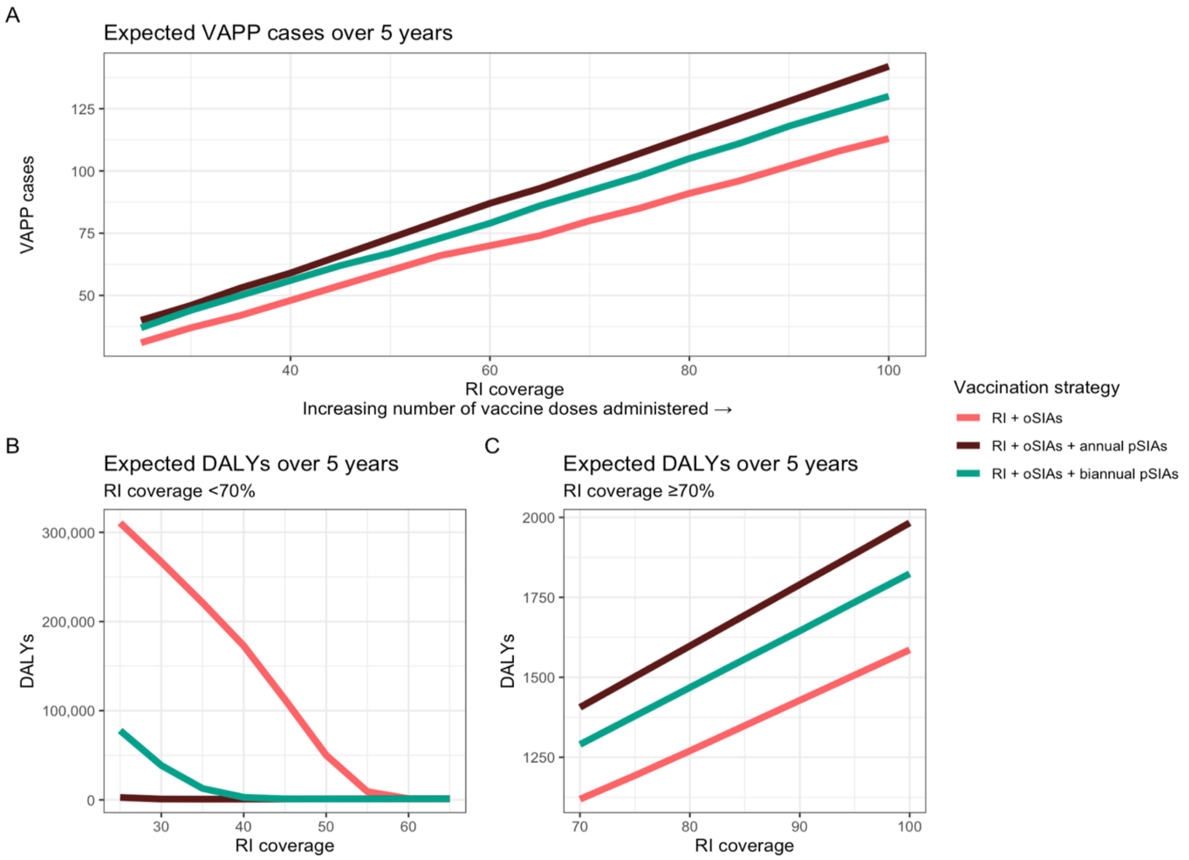


*Appendix Figure 4. Average number of expected VAPP cases and DALYs per capita over 5 years amongst model simulations that reported at least one outbreak. The increase in VAPP cases is related to the increase in vaccine doses administered. In figure 4A, the mean number of VAPP cases were obtained from all model simulations for each vaccination strategy. In figures 4B and 4C, DALYs include DALYs associated with both paralytic polio cases and DALYS and are split by two thresholds of RI coverage, above and below 70%, in order to illustrate the reversal in strategy ranking for DALYs at low and high levels of RI coverage.*

### Section 4: Sensitivity analyses

#### Sensitivity analysis—discounting for health

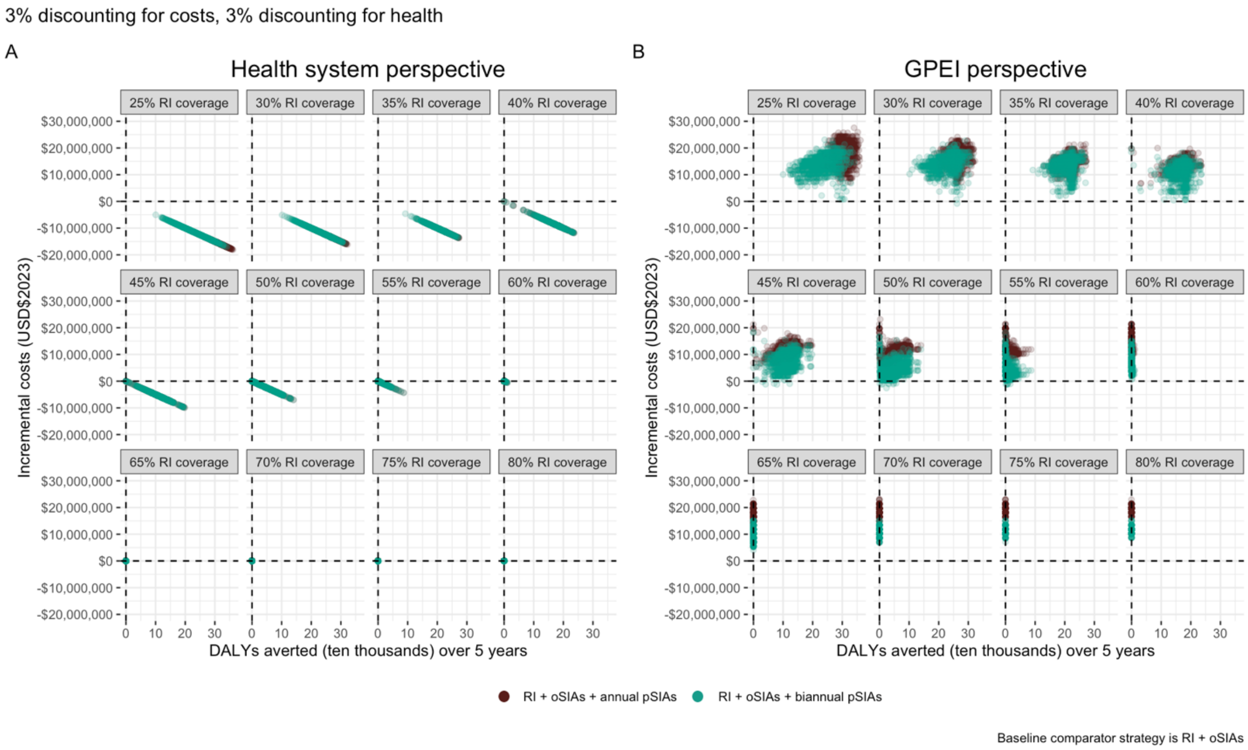


*Appendix Figure 5. Incremental costs per DALY averted under the annual pSIA and biannual pSIA strategies in comparison to the RI+oSIA strategy assuming a 3% discount rate for costs and 3% discount rate for health. Incremental costs are split between health system/non-GPEI perspective and GPEI perspective and the plots are facetted by RI coverage level. The points correspond to 1,000 stochastic model simulations.*

#### Sensitivity analysis – different assumptions about vaccine wastage

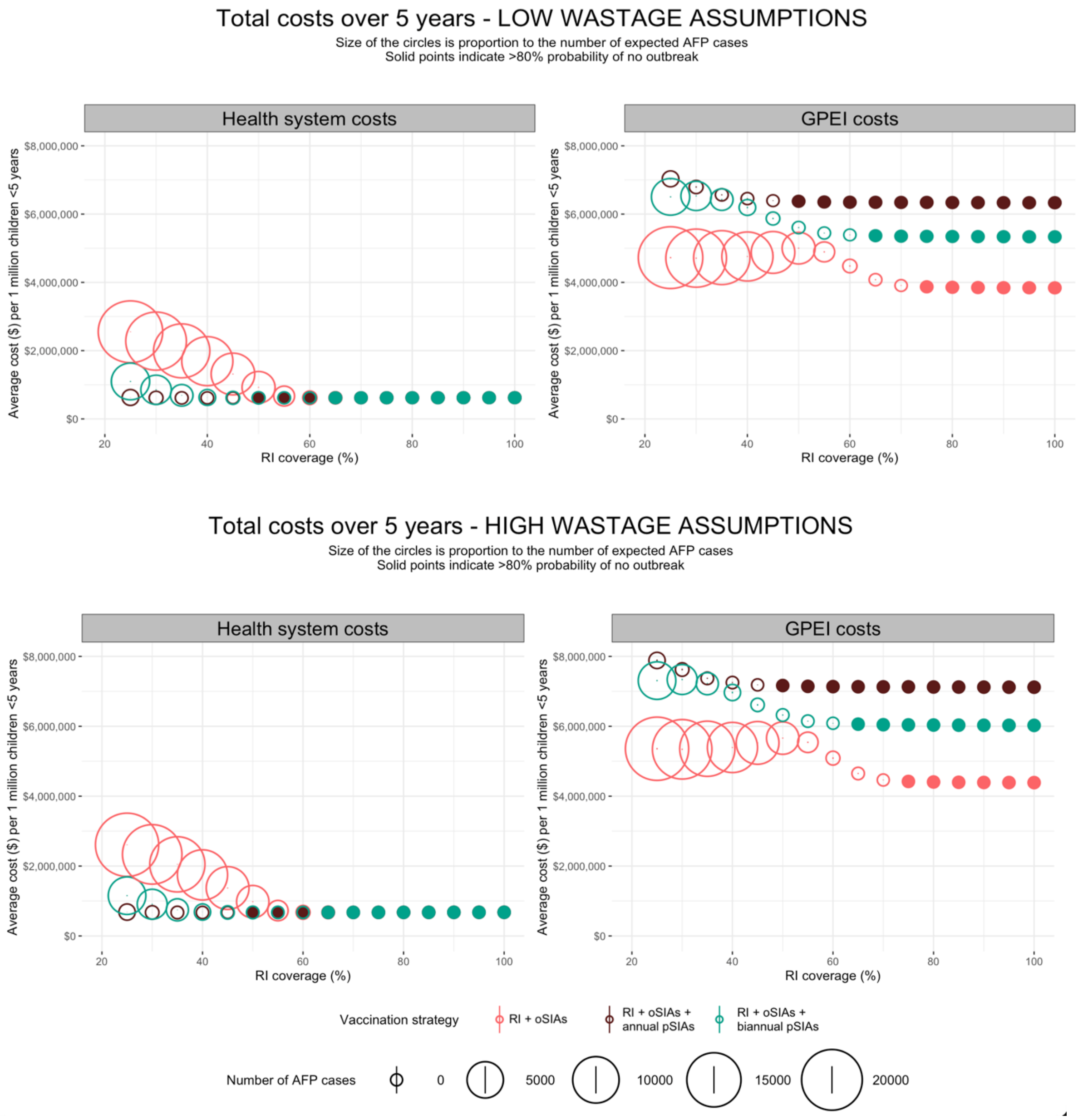


*Appendix Figure 6. Total costs over five years under different wastage assumptions. The top plots represent conservative estimates of wastage using the lowest limit of the range of published wastage rates for both OPV and IPV vaccination via RI and SIAs while the bottom plots use the highest limit of the range of published wastage rates.*

##### Sensitivity analysis – different proportional costs between pSIAs and oSIAs

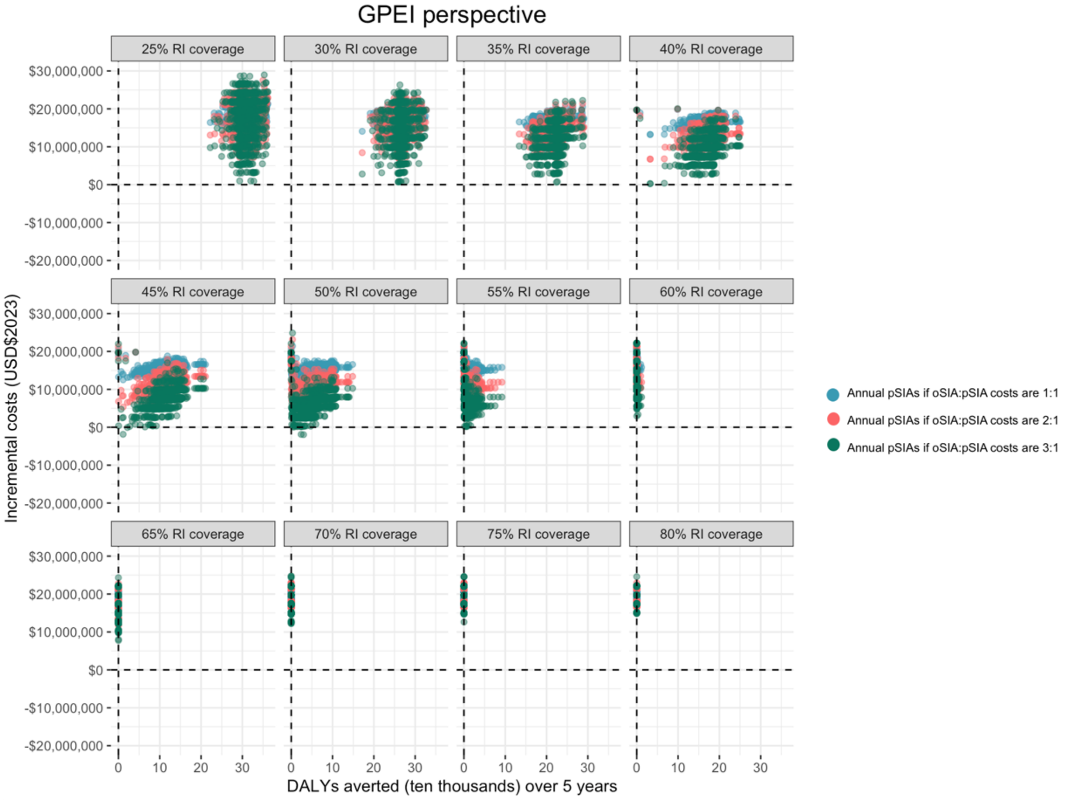


*Appendix Figure 7. Sensitivity analysis exploring incremental costs per DALY averted under variable assumptions about oSIA costs. The different colours represent different proportional differences between annual pSIAs and oSIAs. For example, "Annual pSIAs if oSIA:pSIA costs are 3:1" presents the incremental costs and DALYs averted by annual pSIAs if oSIAs cost three times as much as pSIAs. The points correspond to 1,000 stochastic model simulations.*

A sensitivity analysis explored incremental costs under variable assumptions about the proportional difference in costs between oSIAs and annual pSIAs. The main analysis assumed the cost per child for an oSIA was twice the cost per child for a pSIA, but available outbreak response cost data suggests a range across African countries. In the sensitivity analysis, two alternative cost assumptions were explored: (I) the cost per child in an oSIA equalled the cost per child in a pSIA (1:1) and (II) the cost per child of an oSIA was three times the cost per child in a pSIA (3:1). The RI+oSIA strategy is always the baseline comparator, but here, the total costs of the baseline comparator change: (blue) baseline comparator is oSIAs if the proportional costs are 1:1 with pSIAs, (pink) baseline comparator is oSIAs if the proportional costs are 2:1 (as in the main analysis), and (green) baseline comparator is oSIAs if the proportional costs are 3:1 with pSIAs. This sensitivity analysis answers the question, under what cost assumptions are annual pSIAs more cost-effective than oSIAs? Appendix Figure 7 shows the incremental costs per DALY averted across all oSIA assumptions and Appendix Figure 8 shows incremental costs per outbreak averted from the GPEI perspective as GPEI costs include SIA costs. When RI coverage exceeds the herd immunity threshold, the number of outbreaks, and consequently, the number of oSIAs required substantially decreases causing the GPEI incremental costs to be aligned across all oSIA cost assumptions. Using the equation for Incremental Cost Effectiveness Ratios (ICER), we can comment on the correlation between different proportional costs and the change of direction in incremental costs:

$$ICER = \frac{(costs\ of\ pSIA\ strategy - costs\ of\ RI + oSIA\ strategy)}{(DALYs\ or\ outbreaks\ averted\ by\ pSIA\ strategy)}$$

If we focus on DALYs, when RI coverage is below 60%, annual pSIAs are the least cost-effective when proportional costs between pSIAs and oSIAs are 1:1 because under this assumption, the RI+oSIA strategy is less costly, so the numerator increases. Or, the annual pSIA strategy costs much more than the oSIA per DALY averted. Conversely, when proportional costs between pSIAs and oSIAs are 3:1, the numerator shrinks, so the difference in costs between annual pSIAs and oSIAs is less per DALY averted. When RI coverage increases above 60%, few to no DALYs are averted by annual pSIAs in comparison to oSIAs, but costs remain high as the annual pSIA strategy maintains annual campaigns regardless of outbreak number.

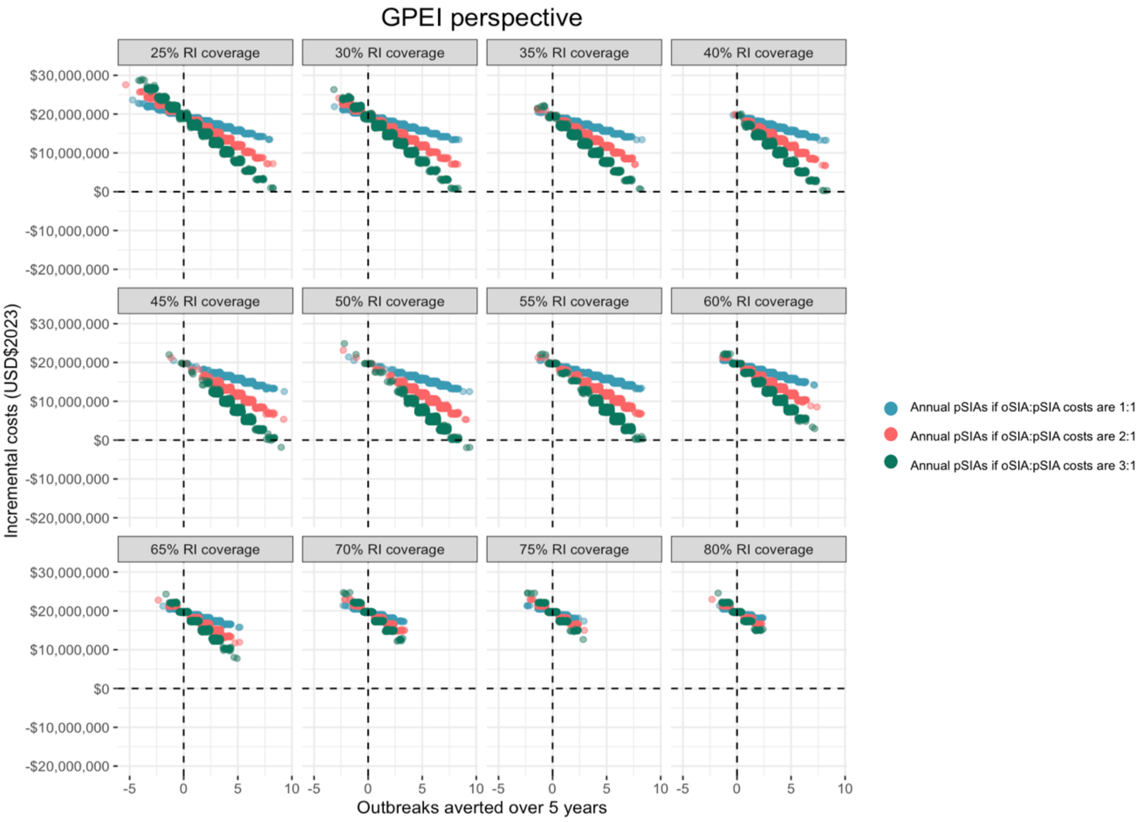


*Appendix Figure 8. Sensitivity analysis exploring incremental costs per outbreak averted under variable assumptions about oSIA costs. The different colours represent different proportional differences between pSIAs and oSIAs. For example, "Annual pSIAs if oSIA:pSIA costs are 3:1" presents the incremental costs and outbreaks averted by annual pSIAs if oSIAs cost three times as much as pSIAs. The points correspond to 1,000 stochastic model simulations.*

Focussing on outbreaks, the annual pSIA strategy does not avert any outbreaks when RI coverage is below 40%, a phenomenon described in the main text for figure 6. At these RI coverage levels, the annual pSIA strategy costs more to avert no outbreaks, and if annual pSIAs are the same cost as oSIAs, the incremental costs are even high per outbreak (not) averted. When RI coverage exceeds 40%, the annual pSIA strategy averts outbreaks but is more costly than the RI+oSIA strategy. Specifically, if the proportional costs are 1:1, the incremental costs of annual pSIAs per outbreak averted are greatest and least when proportional costs between pSIAs and oSIAs are 3:1.

#### Sensitivity analysis – different assumptions about proportion of children reached by SIAs

The true effectiveness of SIAs (defined as the product of coverage and vaccine efficacy) remains uncertain in practice; although vaccine efficacy is well described in clinical trials, it is known to vary by population, and the population coverage achieved is uncertain resulting in variable effectiveness. For example, mathematical modelling demonstrates a dramatic range of SIA effectiveness estimates in Tajikistan versus the Republic of Congo: in Tajikistan, SIA effectiveness in response to a localised outbreak was estimated to be 69% (95% CI 55-80%) while in the Republic of Congo, SIA effectiveness was estimated to be 0.4% (95% CI 0.0-14.0%) per SIA [11]. In this analysis we define the SIA target population as children missed by RI. As research suggests that SIA effectiveness is highly variable in different locations, this model assessed outcomes under the assumption that all pSIAs and oSIAs reach 25% of the population missed by RI and bOPV vaccine efficacy is 50%. In the sensitivity analysis below (Appendix Table 4), we explore the average number of expected AFP cases over five years and the probability of an outbreak if SIAs reached 50% of the target population, or 50% of children under five years of age who were missed by RI.

*Appendix Table 4. Expected average number of AFP cases and expected probability of an outbreak if each SIA were expected to reach 50% of the target population instead of the 25% assumption in the main analysis. Assumptions about vaccine efficacy are the same across all models. Expected AFP cases were calculated by taking the average number of AFP cases across all stochastic model simulations that had **at least one AFP case.***

| RI coverage | SIA strategy | If SIAs vaccinate 25% of the target population | | If SIAs vaccinate 50% of the target population | |
|---|---|---|---|---|---|
| | | Average number of expected AFP cases over 5 years | Expected probability of an outbreak | Average number of expected AFP cases over 5 years | Expected probability of an outbreak |
| 25% | RI + oSIA + annual pSIA | 176 | 88% | 1 | 16% |
| 35% | RI + oSIA + annual pSIA | 5 | 65% | 1 | 9% |
| 50% | RI + oSIA + annual pSIA | 1 | 20% | 1 | 6% |
| 75% | RI + oSIA + annual pSIA | 1 | 4% | 1 | 2% |
| 25% | RI + oSIA + biannual pSIA | 5,564 | 99% | 15 | 83% |
| 35% | RI + oSIA + biannual pSIA | 894 | 97% | 3 | 56% |
| 50% | RI + oSIA + biannual pSIA | 4 | 67% | 1 | 17% |
| 75% | RI + oSIA + biannual pSIA | 1 | 6% | 1 | 6% |
| 25% | RI + oSIA | 22,158 | 100% | 18,093 | 100% |
| 35% | RI + oSIA | 15,753 | 100% | 9,851 | 100% |
| 50% | RI + oSIA | 3,549 | 99% | 602 | 99% |
| 75% | RI + oSIA | 1 | 14% | 1 | 14% |

#### Sensitivity analysis – different $R_0$ assumptions

As the true value of $R_0$ for polio is unknown and depends on geographical settings, sanitation and hygiene, and age. In the main analysis, we assume an $R_0$ of 3, which is in line with other research and the target population of children under five years of age living in an LMIC in Africa. However, in the sensitivity analysis below (Appendix Table 5) we explore the average number of expected AFP cases over five years and the probability of an outbreak if $R_0$ = 6. As shown in Appendix Table 5, when RI coverage >50%, a higher $R_0$ only increases the probability of an outbreak slightly in the annual pSIA strategy but has a bigger effect on outbreak probability for the biannual pSIA and RI+oSIA strategies.

*Appendix Table 5. Expected number of AFP cases and expected probability of an outbreak under different $R_0$ assumptions. Expected AFP cases were calculated by taking the average number of AFP cases across all stochastic model simulations that had* ***at least one AFP case.***

| RI coverage | SIA strategy | $R_0$ = 3 | | $R_0$ = 6 | |
|---|---|---|---|---|---|
| | | Average number of expected AFP cases over 5 years | Expected probability of an outbreak | Average number of expected AFP cases over 5 years | Expected probability of an outbreak |
| 25% | RI + oSIA + annual pSIA | 176 | 88% | 16,379 | 100% |
| 50% | RI + oSIA + annual pSIA | 1 | 20% | 3,837 | 100% |
| 75% | RI + oSIA + annual pSIA | 1 | 4% | 1 | 19% |
| 25% | RI + oSIA + biannual pSIA | 5,564 | 99% | 24,403 | 100% |
| 50% | RI + oSIA + biannual pSIA | 4 | 67% | 9,542 | 100% |
| 75% | RI + oSIA + biannual pSIA | 1 | 6% | 4 | 68% |
| 25% | RI + oSIA | 22,158 | 100% | 37,069 | 100% |
| 50% | RI + oSIA | 3,549 | 99% | 20,229 | 100% |
| 75% | RI + oSIA | 1 | 14% | 2,046 | 99% |

#### Sensitivity analysis – different importation rates

When RI coverage exceeds 50%, model assumptions for the importation rate of WPV infection have little effect on the expected number of AFP cases and expected probability of an outbreak across all vaccination strategies. This reiterates the importance of baseline RI coverage, which remains an important underlying factor that greatly influences the number of expected AFP cases and outbreak probability, more so than importation rate or assumptions around the proportion of children reached by SIAs.

*Appendix Table 6. Expected number of AFP cases and expected probability of an outbreak if the rate of WPV importation is one importation of WPV infection every year and three importations every year, rates that are under and over the rate used in the main analysis (two importations per year). Expected AFP cases were calculated by taking the average number of AFP cases across all stochastic model simulations that had* ***at least one AFP case.***

| RI coverage | SIA strategy | 1 importation every year | | 3 importations every year | |
|---|---|---|---|---|---|
| | | Average number of expected AFP cases over 5 years | Expected probability of an outbreak | Average number of expected AFP cases over 5 years | Expected probability of an outbreak |
| 25% | RI + oSIA + annual pSIA | 42 | 71% | 49 | 96% |
| 50% | RI + oSIA + annual pSIA | 1 | 11% | 1 | 27% |
| 75% | RI + oSIA + annual pSIA | 1 | 2% | 1 | 5% |
| 25% | RI + oSIA + biannual pSIA | 5,353 | 95% | 5,363 | 100% |
| 50% | RI + oSIA + biannual pSIA | 4 | 45% | 5 | 78% |
| 75% | RI + oSIA + biannual pSIA | 1 | 3% | 1 | 8% |
| 25% | RI + oSIA | 22,327 | 99% | 22,236 | 100% |
| 50% | RI + oSIA | 3,430 | 91% | 3,399 | 100% |
| 75% | RI + oSIA | 1 | 9% | 1 | 20% |

### Section 5: Supporting information

#### Raw data used to inform proportional differences between pSIA and oSIA costs

*Appendix Table 7. Country level SIA data showing the differences in costs between oSIAs and pSIAs. The rightmost column shows the proportional difference between oSIAs and pSIAs for operational costs. *pSIA cost data was available pre COVID-19 pandemic and given in USD$2019, therefore, estimates have been calculated for USD$2023 assuming 2019$1 = 2023$1.18. oSIA cost data for bOPV has historically been less readily available. Therefore, to estimate the proportional differences between oSIAs and pSIAs, oSIA cost data from campaigns administering novel OPV2 (nOPV2) was used and we assume that while the vaccine costs may differ, the oSIA operational costs are similar.*

| Country | bOPV cost (USD$) | pSIA - cost per child $2019 | pSIA - cost per child $2023* | oSIA (nOPV) - cost per child $2023 | proportional difference - *operational only* |
|---|---|---|---|---|---|
| Benin | 0.17 | 0.31 | 0.37 | 0.73 | 2.0 |
| Burkina Faso | 0.15 | 0.27 | 0.32 | 0.65 | 2.0 |
| Cameroon | 0.17 | 0.33 | 0.39 | 0.86 | 2.2 |
| Central African Republic | 0.17 | 1.09 | 1.29 | 2.79 | 2.2 |
| Chad | 0.16 | 0.45 | 0.53 | 0.42 | 0.8 |
| Congo | 0.16 | 0.44 | 0.52 | 2.29 | 4.4 |
| Côte d'Ivoire | 0.14 | 0.15 | 0.18 | 0.67 | 3.8 |
| DR Congo | 0.15 | 0.48 | 0.57 | 1.28 | 2.3 |
| Eritrea | 0.18 | 0.95 | 1.12 | 1.00 | 0.9 |
| Ethiopia | 0.17 | 0.89 | 1.05 | 0.56 | 0.5 |
| Gabon | 0.16 | 0.77 | 0.91 | 1.00 | 1.1 |
| Gambia | 0.18 | 0.60 | 0.71 | 0.78 | 1.1 |
| Ghana | 0.18 | 0.29 | 0.34 | 1.32 | 3.9 |
| Guinea | 0.19 | 0.26 | 0.31 | 1.23 | 4.0 |
| Kenya | 0.18 | 0.59 | 0.70 | 1.02 | 1.5 |
| Liberia | 0.20 | 0.83 | 0.98 | 1.58 | 1.6 |
| Madagascar | 0.17 | 0.37 | 0.44 | 0.53 | 1.2 |
| Malawi | - | - | 0.00 | 1.00 | |
| Mali | 0.17 | 0.24 | 0.28 | 0.52 | 1.8 |
| Mauritania | 0.12 | 0.81 | 0.96 | 1.00 | 1.0 |
| Mozambique | - | - | 0.00 | 0.80 | |
| Niger | 0.19 | 0.36 | 0.42 | 0.57 | 1.3 |
| Nigeria | 0.21 | 0.32 | 0.38 | 0.22 | 0.6 |
| Senegal | 0.18 | 0.29 | 0.34 | 0.47 | 1.4 |
| Sierra Leone | 0.16 | 0.46 | 0.54 | 0.92 | 1.7 |
| South Sudan | 0.17 | 0.78 | 0.92 | 1.17 | 1.3 |
| Sudan | 0.15 | 0.58 | 0.68 | 0.65 | 0.9 |
| Tanzania | 0.17 | 0.93 | 1.10 | 1.00 | 0.9 |
| Togo | 0.17 | 0.27 | 0.32 | 0.73 | 2.3 |
| Uganda | 0.18 | 0.48 | 0.57 | 1.00 | 1.8 |
| Zambia | - | - | | 2.78 | |
| Zimbabwe | - | - | | 1.76 | |
| **Average** | **$0.17** | **$0.52** | **$0.61** | **$1.04** | **$1.7** |

#### Cost-effectiveness plane to interpret incremental costs discussed in main text

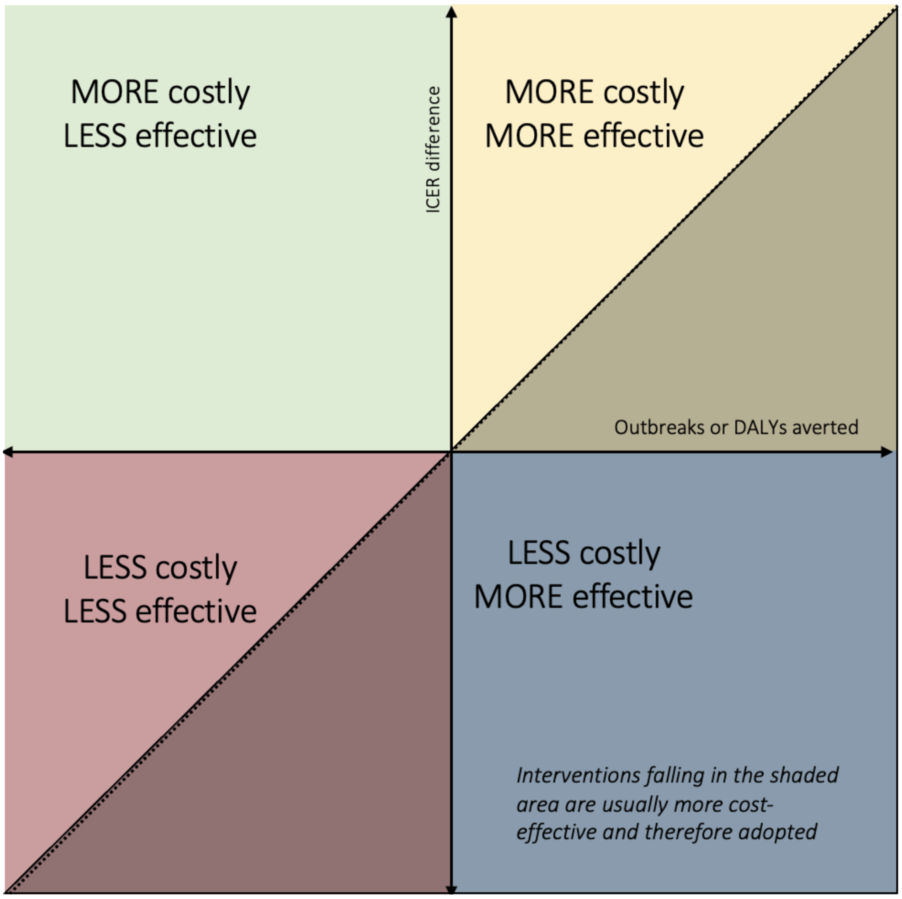


*Appendix Figure 9. Quadrants comprising a cost effectiveness plane for interpretation of incremental costs and DALYs and outbreaks averted under each vaccination strategy.*
